# Discrepancy review: A feasibility study of a novel peer review intervention to reduce undisclosed discrepancies between registrations and publications

**DOI:** 10.1101/2022.01.18.22269507

**Authors:** TARG Meta-Research Group & Collaborators, Robert T. Thibault, Tom E. Hardwicke, Robbie W. A. Clark, Charlotte R. Pennington, Gustav Nilsonne, Aoife O'Mahony, Katie Drax, Jacqueline Thompson, Marcus R. Munafò

## Abstract

**Background:** Undisclosed discrepancies often exist between study registrations and their associated publications. Discrepancies can increase risk of bias, and when undisclosed, they disguise this increased risk of bias from readers. To remedy this issue, we developed an intervention called *discrepancy review*. We provided journals with peer reviewers specifically assigned to check for undisclosed discrepancies between registrations and manuscripts submitted to journals.

**Objectives:** We aimed to (1) evaluate the feasibility of incorporating discrepancy review as a regular practice at scientific journals and the feasibility of conducting a trial on discrepancy review; (2) explore the benefits and time required to incorporate discrepancy review as a regular practice at scientific journals; and (3) refine the discrepancy review process.

**Method:** We performed discrepancy review on 18 manuscripts submitted to *Nicotine and Tobacco Research* and 3 manuscripts submitted to the *European Journal of Personality*. We iteratively refined the discrepancy review process based on feedback from discrepancy reviewers, editors, and authors. We then assessed whether revised manuscripts addressed recommendations from discrepancy reviewers and identified potential outcome measures for use in a future trial of discrepancy review.

**Results:** Registrations were generally too imprecise to be effectively evaluated by our original discrepancy review process so we developed a simplified, semi-structured process. Authors addressed the majority of discrepancy reviewer comments and there was no opposition to running a trial from authors, editors, or discrepancy reviewers. Clinical trial registrations were more precise but less comprehensive than registrations on the Open Science Framework, suggesting they should be evaluated in separate trials. Outcome measures for a trial of discrepancy review on clinical trial registration could include the presence of primary or secondary outcome discrepancies and whether publications that are not the primary report from a clinical trial registration are clearly described as such. Outcome measures for a trial on Open Science Framework registrations could include assessments of whether registrations are permanent, as well as an overarching subjective assessment of the impact of discrepancies.

**Conclusion:** We found that discrepancy review could feasibly be introduced as a regular practice at journals interested in this process. A full trial of discrepancy review would be needed to evaluate its impact on reducing undisclosed discrepancies.

## 1. Introduction

Prospective registration of scientific studies serves several purposes and has become standard practice for clinical trials (Zarin et al., 2017). Other research disciplines have followed suit and created their own implementations of prospective registration, often called preregistration. These disciplines include psychology, economics, and other social sciences; systematic reviews and meta-analyses; and to some extent, preclinical animal research and observational health research. Across these disciplines, one of the primary objectives of registering study plans is to clearly demarcate confirmatory research from exploratory research, and in turn, reduce or at least expose, outcome switching, selective reporting, and data dredging (Bakker et al., 2020; Hardwicke & Wagenmakers, 2021; Nosek et al., 2018; Zarin et al., 2017).

In practice, published studies often contain undisclosed discrepancies in relation to their registration. A meta-analysis of more than 6,000 publications and their associated registrations, mostly in clinical medicine, estimated that 10-68% of registered studies have at least one primary outcome discrepancy and 13-95% have at least one secondary outcome discrepancy (95% prediction intervals) (TARG Meta-Research Group & Collaborators, 2021). Undisclosed discrepancies are also common in psychology (Claesen et al., 2021) and economics research (Ofosu & Posner, 2019). While a publicly available registration allows readers to check whether the outcomes reported in a publication match those that were initially planned, it would be unrealistic to expect every reader to examine this information. Indeed, one survey suggested that only about a third of clinical trial peer reviewers examine registrations as part of their review (Mathieu et al., 2013). Moreover, if a reader finds discrepancies between a registration and publication, there is no clear avenue for how to share this information. One study systematically sent 58 letters to the editor that described undisclosed discrepancies in published manuscripts and found that only three of five top medical journals were willing to publish these letters (Goldacre et al., 2019). A publication workflow that addresses reporting issues *before* publication would be more desirable than relying on individual readers’ scrutiny and correction efforts. One ongoing trial attempts to reduce discrepancies at medical journals by sending peer reviewers information about the registration associated with the manuscript they are reviewing (protocol available at: Jones et al., 2019). This ongoing study leaves the decision for how to use this information to the reviewers and editors.

Here, we propose a novel intervention—*discrepancy review—*to improve transparent reporting between registrations and their final manuscript. Our intervention provides journals with peer reviewers specifically assigned to check for both outcome and non-outcome discrepancies and asks them to prepare an itemized list of constructive recommendations to manuscript authors for how to reduce or disclose discrepancies between their registration and submitted manuscript. Importantly, the goal of discrepancy review is to encourage transparent reporting, not to inhibit publication of manuscripts with discrepancies. In many cases, discrepancies are entirely justifiable and can be necessary to improve a study design or analysis.

### 1.1 Objectives

We preregistered three overarching objectives, but no hypotheses. Our objectives were to: (1a) evaluate the feasibility of incorporating discrepancy review as a regular practice at scientific journals, (1b) evaluate the feasibility of conducting a trial on discrepancy review; (2) explore the benefits and effort required to incorporate discrepancy review as a regular practice at scientific journals; and (3) refine the discrepancy review process.

### 1.2 Terminology

We use the term *registration* throughout this manuscript to refer to time-stamped documents stored in a permanent and publicly accessible repository (e.g., clinicaltrials.gov, osf.io/registries) that contain information about a study (e.g., design, outcomes, analyses). Registrations can be *prospective* (posted before participant enrollment) or *retrospective* (after participant enrollment begins). Another term for prospective registration is *preregistration*. While these terms are sometimes used interchangeably, clinical trials often use the term prospective registration, and various other disciplines often use the term preregistration. Clinical trial registrations differ from registration on platforms such as the Open Science Framework (OSF) in their history, the functions they were designed to serve, and implementation details (e.g., clinical trial registries do not include sections dedicated to hypotheses or analysis plans). Because of these differences, we try not to conflate clinical trial registration with preregistration more broadly and mostly use the term registration, which encompasses both, and makes no assumption regarding the timing of registration.

We use the term *discrepancy* to refer to any incongruity between the content of a manuscript and its associated registration. We use the term *discrepancy review* to encompass the entire process we developed to check for and report on discrepancies, and *discrepancy report* for the written report that comes from discrepancy review and is shared with the action editor.

## 2. Methods

### 2.1 Journal recruitment

Via email, we invited the editors-in-chief of 18 journals in medicine and psychology to participate. We selected 11 journals from the list that offer preregistered badges (cos.io/initiatives/badges) as well as 7 additional journals our team thought might be interested in participating.

### 2.2 Discrepancy reviewers

Discrepancy reviewers included the project lead, a frequent collaborator, and three team members recruited from a call on Twitter and selected for their experience with registration and range of career stages. While all the reviewers have experience with meta-research and registration, as well as general knowledge in neuroscience or psychology, they did not have domain expertise in the topics of personality psychology and nicotine/tobacco research which they reviewed. At the time of reviewing, their career stages included associate professor (GN, *n* discrepancy reviews performed = 3), assistant professor (CRP, *n* = 10), postdoctoral researcher (RTT, *n* = 21; TEH, *n* = 5), and PhD student (AO, *n* = 3).

### 2.3 Original discrepancy review

Before starting this feasibility study, we developed a systematic peer review procedure—discrepancy review—to detect discrepancies between registrations and submitted manuscripts across the 18 dimensions listed in Table 1. To perform discrepancy review, a team member would complete a survey regarding how these 18 dimensions were reported in the registration^1^, repeat the process using the submitted manuscript, and compare the output of the surveys. They would identify the presence of discrepancies across each dimension as well as whether the discrepancies were disclosed, and provide a subjective assessment of whether each discrepancy presents an issue that they considered negligible, minor, or major. The reviewer would then prepare a *discrepancy report* that itemizes the discrepancies and provide recommendations for how the authors can address them. The reviewer would submit this report to the action editor, who then sends it to the manuscript authors.

**Table 1.** Reporting and discrepancies across the 18 dimensions included in the original discrepancy review process. Reporting was coded as 0 (not reported), 1 (reported, but unclear), or 2 (reported clearly). This table presents data from the first coder for the 10 manuscripts^5^ that underwent the original discrepancy review process and were not a secondary publication associated with a clinical trial registration^6^.

| Dimension | Registration |  |  | Manuscript |  |  | Discrepancy |  |  |  | Issues |  |  |  |  |
| --- | --- | --- | --- | --- | --- | --- | --- | --- | --- | --- | --- | --- | --- | --- | --- |
|  | 0 | 1 | 2 | 0 | 1 | 2 | NA | no | yes | disclosed | NA | none | negligible | minor | major |
| Hypotheses (primary) | 0 | 4 | 6 | 2 | 5 | 3 | 0 | 5 | 5 | 1 | 0 | 5 | 0 | 1 | 4 |
| Hypotheses (secondary) | 7 | 3 | 0 | 8 | 2 | 0 | 6 | 0 | 4 | 0 | 6 | 1 | 0 | 2 | 1 |
| Independent variables | 0 | 2 | 8 | 0 | 2 | 8 | 0 | 9 | 1 | 0 | 0 | 7 | 2 | 0 | 1 |
| Covariates / moderators | 7 | 2 | 1 | 6 | 2 | 2 | 6 | 2 | 2 | 0 | 4 | 3 | 1 | 1 | 1 |
| Outcomes (primary) | 0 | 3 | 7 | 0 | 2 | 8 | 0 | 5 | 5 | 0 | 0 | 4 | 0 | 2 | 4 |
| Outcomes (secondary) | 4 | 4 | 2 | 6 | 2 | 2 | 3 | 2 | 5 | 0 | 3 | 2 | 0 | 1 | 4 |
| Sample size | 1 | 1 | 8 | 1 | 0 | 9 | 1 | 2 | 7 | 0 | 1 | 2 | 3 | 1 | 3 |
| Sample size justification | 6 | 1 | 3 | 10 | 0 | 0 | 6 | 0 | 4 | 0 | 6 | 0 | 0 | 2 | 2 |
| Participant eligibility | 3 | 0 | 7 | 2 | 0 | 8 | 1 | 3 | 6 | 0 | 0 | 4 | 4 | 2 | 0 |
| Data exclusion | 6 | 3 | 1 | 7 | 1 | 2 | 6 | 1 | 3 | 0 | 4 | 0 | 3 | 3 | 0 |
| Missing data | 8 | 1 | 1 | 5 | 1 | 4 | 5 | 1 | 4 | 1 | 1 | 1 | 7 | 1 | 0 |
| Randomization | 5 | 3 | 2 | 5 | 1 | 4 | 5 | 5 | 0 | 0 | 5 | 3 | 2 | 0 | 0 |
| Blinding | 6 | 2 | 2 | 8 | 1 | 1 | 6 | 1 | 3 | 0 | 5 | 1 | 1 | 2 | 1 |
| Preprocessing | 10 | 0 | 0 | 9 | 1 | 0 | 9 | 0 | 1 | 0 | 7 | 1 | 2 | 0 | 0 |
| Analysis (primary) | 2 | 5 | 3 | 0 | 4 | 6 | 0 | 1 | 9 | 0 | 0 | 2 | 0 | 5 | 3 |
| Statistical assumptions | 8 | 2 | 0 | 9 | 1 | 0 | 7 | 0 | 3 | 0 | 5 | 0 | 3 | 2 | 0 |
| Inferential criteria | 6 | 0 | 4 | 7 | 1 | 2 | 5 | 2 | 3 | 0 | 4 | 1 | 4 | 0 | 1 |
| Analysis (secondary) | 4 | 5 | 1 | 8 | 1 | 1 | 4 | 0 | 6 | 1 | 4 | 2 | 1 | 3 | 0 |

Supplementary Material C operationalizes each dimension. Supplementary Table B2 contains the same data from a second coder on the subset of manuscripts where the second coder also performed the original discrepancy review process. We did not attempt to resolve differences between coders. Inter-rater agreement is available in Supplementary Table B3. Issues were considered NA if the dimension was irrelevant to the particular study (e.g., if neither the registration nor manuscript had secondary hypotheses). Issues were coded as present for some dimensions due to a lack of transparency rather than a discrepancy.

### 2.4 Updated discrepancy review

Due to shortcomings in the original discrepancy review process (see Section 3.2), which we used on 16 manuscripts, we developed and switched to a simplified process (summarized in Box 1). Whereas the original discrepancy review process used a detailed and structured checklist, the updated discrepancy review process had a semi-structured format with guiding questions. These changes aimed to reduce the time required to perform discrepancy review, while still identifying all important discrepancies.

#### Box 1.

Summary of the eight questions guiding the updated discrepancy review process.

1. Does the manuscript present only **exploratory** outcomes and analyses?
2. Is the registration **properly registered** (i.e., permanent and public)?
3. Does the registration date suggest that the timing of registration may be **retrospective**?
4. Are there discrepancies in the number, content, or prioritization of **hypotheses**?
5. Do the study arms, **independent variables**, exposure variables, or study/experimental grouping match?*
6. Are there discrepancies in the number, content, or prioritization of **outcome measures**?
7. Do the **analyses** match?*
8. While answering the previous questions, if you identified notable **additional discrepancies** or questionable research practices you may raise them. These could include sample size, control variables, covariates or moderators, eligibility criteria, analytic decisions (e.g., outlier definition), randomization, blinding, and data preprocessing, among others.*

*For these items, reviewers were instructed to not spend time looking for minor discrepancies. Supplementary Material E contains the exact instructions for the updated discrepancy review process.

### 2.5 Discrepancy reports

Two team members independently prepared a written discrepancy report for each manuscript. Only the first report was sent to the action editors. We used the second report to examine consistencies and differences among discrepancy reviewer reports. For eight manuscripts, both the first and second discrepancy reports were written after performing the original discrepancy review process; for eight manuscripts, the first report was based on the original discrepancy review process and the second report was based on the updated discrepancy review process; for five manuscripts, both reports were based on the updated discrepancy review process.

## 3. Results

### 3.1 Journal recruitment

Of the 18 journals we contacted, five agreed to participate, two were interested in participating but did not begin sending us manuscripts by the time we concluded data collection, six declined to participate, and five did not respond. Of the five journals that agreed to participate, two did not receive any manuscripts reporting a registration during the study period, and one had difficulty adding discrepancy review to their manuscript handling procedures and therefore did not send us any manuscripts to review.

Of the journals that declined to participate, three expressed interest but were currently too busy, one stated they receive too few manuscripts reporting a registration and believed the process would be complicated to set up, one stated they were uncomfortable using their journal for research purposes, and one stated their editors already check for discrepancies rendering their journal ill-suited for the present study. All but two invitations were sent within the first eight months of the Covid-19 pandemic, which may have impacted the willingness of journals to participate.

We reviewed 18 manuscripts submitted to *Nicotine and Tobacco Research* over a period of three months and 3 manuscripts submitted to the *European Journal of Personality* over a period of five months. When assigning manuscripts to peer review, the action editors at *Nicotine and Tobacco Research* additionally invited our team to perform discrepancy review. These reviews were submitted through an online submission portal (ScholarOne) within three weeks of invitation, as done for regular peer reviews. One of our team members (MRM) is the editor-in-chief of *Nicotine and Tobacco Research* and served as action editor on two manuscripts included in this study, but did not perform any of the discrepancy reviews. The editor-in-chief of the *European Journal of Personality* invited our team to review manuscripts on which he himself served as the action editor. For manuscripts that passed an initial round of review at the *European Journal of Personality*, the editor-in-chief shared the manuscript with our team and informed authors that their manuscript would soon undergo discrepancy review. We conducted discrepancy review within a week and the editor-in-chief sent our review to the manuscript authors in an email separate from the standard peer reviewer comments and editorial decision.

We suggested that journals inform manuscript authors about the study in one of four ways: (1) an opt-*in* button on the journal’s manuscript submission page; (2) an opt-*out* button on the journal’s manuscript submission page; (3) through an automated email following manuscript submission; or (4) in the email containing the initial editorial decision. One journal chose Option 1 and had an opt-in rate of 79% over four months. The opt-in text was relatively vague and stated that the journal had partnered with researchers to improve peer review, but did not mention anything about registration in particular^2^. Due to difficulties setting up the opt-in button, editors forgetting to flag manuscripts that both opted-in and reported a registration, and a limited number of manuscripts that met both these criteria, we did not receive invitations to review manuscripts from this journal. One participating journal selected Option 4, informing authors in the same email as the initial editorial decision and standard peer review comments. The other participating journal selected a modified version of Option 3, where they manually sent an email to authors after an initial editorial decision to revise and resubmit, but before discrepancy review. These emails informed authors that they could withdraw the data collected based on their manuscript. No author requested their data be withdrawn.

### 3.2 Original discrepancy review

We planned to use discrepancy review for two purposes: to provide the basis for writing discrepancy reports (the intervention), and to evaluate the presence of discrepancies in published manuscripts (for outcome assessment). We found our original implementation of discrepancy review fit for neither purpose; mainly because registrations were less comprehensive and less precise than we anticipated. Supplementary Table B1 lists additional shortcomings in the original discrepancy review process and how we addressed them. Nonetheless, the data collected from the original discrepancy review process, which we used as the basis for writing discrepancy reports, provided several insights (Table 1): many dimensions were rarely included in registrations and manuscripts, such as sample size justifications, plans for data exclusion, secondary hypotheses, and secondary analyses; discrepancies were common across many dimensions; author disclosure of discrepancies was very rare; many discrepancies were judged by reviewers to be negligible; and, most discrepancies judged to be major issues are in hypotheses, outcomes, sample size, and analyses. Based on these findings, we developed an updated discrepancy review process and revised our outcome measures.

#### 3.2.1 Inter-rater agreement: original discrepancy review

Inter-rater agreement was low across many of the 18 dimensions coded in the original discrepancy review process (inter-rater comparisons of the written discrepancy *reports* are provided in section 3.4 of this manuscript). In particular, inter-rater agreement was low for judgements of discrepancies as negligible, minor, or major (see Supplementary Table B2 for second coder results and Supplementary Table B3 for inter-rater agreement). The original discrepancy review process allowed for subjectivity. For example, the survey asked reviewers to state whether each of the 18 dimensions was reported ‘sufficiently’ for the reviewer to replicate the study if they had the expertise^3^. We also asked reviewers to judge whether a discrepancy or lack of transparency for each dimension presented a negligible, minor, or major issue^4^. Whereas some reviewers made use of ‘negligible’ for issues of transparency (where an item was reported in neither the registration nor the manuscript), other reviewers often considered these minor issues. Some reviewers also used the option NA (not applicable) more frequently than others, in turn lowering inter-rater agreement. Taken together, we concluded that our original implementation of discrepancy review could not reliably assess whether discrepancies were present, or the severity of their impact.

### 3.3 Updated discrepancy review

Discrepancy reviewers reported preferring the updated process but noted that the less structured methodology required greater focus. The updated discrepancy review process took less time than the original discrepancy review process (see Table 2). We feel updated discrepancy review is more feasible for journals to implement as a standard practice. For two of five manuscripts that underwent updated discrepancy review, and for one of 16 manuscripts that underwent original discrepancy review, the editors invited the discrepancy reviewer for a second round of review to ensure their comments were addressed, as is common for traditional peer review. The times listed in Table 2 are only for the first round of discrepancy review.

**Table 2.**
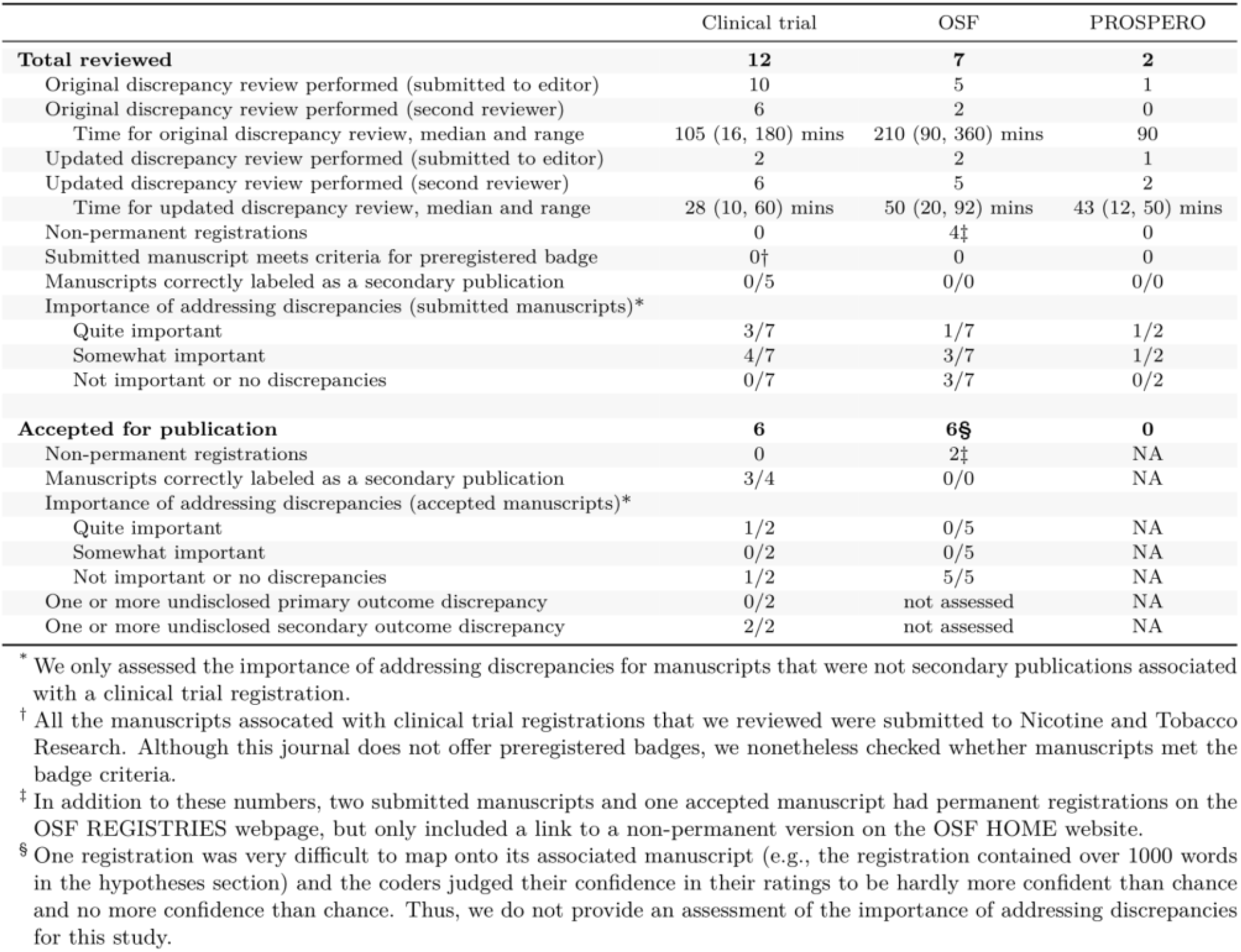
Characteristics and outcomes of the manuscripts reviewed.

### 3.4 Consistency of written discrepancy reports among reviewers

We qualitatively compared each pair of written discrepancy reports and found that their content was relatively similar in regards to substantial discrepancies, but less similar for minor concerns. These were comparisons of the written text in the discrepancy reports and are distinct from the inter-rater agreement we calculated across the 18 dimensions in the original discrepancy review process and the inter-rater agreement of potential outcome measures presented in section 3.6. Regardless of whether reviewers used the original or updated discrepancy review process, their reports were relatively consistent in identifying non-permanent registrations (e.g., study protocols that were publicly shared on an Open Science Framework project page, but not formally registered)^7^; manuscripts that were not the main paper related to a clinical trial registration; substantial discrepancies in the outcome measures of studies that used clinical trial registrations; and substantial discrepancies in the hypotheses of studies using OSF registrations. Three pairs of reports differed considerably. One reviewer reported rushing two reviews and agreed with the presence of major issues when the other reviewer presented them. The third pair of reports were in regards to the first manuscript we received. It was the first manuscript reviewed, but the last manuscript to receive a second review—by which point we had substantially refined how we performed discrepancy review.

Reports often differed in the minor issues they presented, which may have arisen due to the latitude reviewers had in what to report. Reports based on the updated discrepancy review process mentioned fewer minor issues. The style and wording of reports differed somewhat between reviewers. In summary, although inter-rater agreement was low across the specific 18 dimensions coded in the original discrepancy review process, the written reports had comparable overall conclusions.

### 3.5 Editor and author feedback

Five of the 13 action editors involved in our study responded to an optional questionnaire in regards to one of the manuscripts they handled. All five editors responded that they would use discrepancy review again and that it would be possible to implement discrepancy review for all manuscripts submitted to their journal if they were provided with discrepancy reviewers. To limit the impact our study had on the publication process beyond transparent reporting, we specifically asked editors to not allow discrepancy review to influence their editorial decision. We found that four of the five editors reported that discrepancy review did not influence their editorial decision and the fifth reported it encouraged acceptance. Three editors rejected the manuscripts they received and two asked for revisions. Our questionnaire prompted these two editors who asked for revisions with additional questions, such as the time it took them to implement discrepancy review.. They reported taking 6 minutes and 30 minutes and that it was worth their time.

Authors from 4 of the 21 manuscripts responded to an optional questionnaire. They all agreed that they would like their next manuscript to undergo discrepancy review and were positive about discrepancy review in their open-ended responses. They reported taking 30, 90, and 180 minutes to address the discrepancy reviewer comments (one author did not answer this question). Responses from both authors and editors were mixed regarding whether discrepancy review should be implemented before (*n* = 2), during (*n* = 3), or after (*n* = 2) standard peer review (one author responded “unsure” and another author did not respond). The questionnaire included questions about the value of discrepancy review as well as an optional open-ended question asking what they disliked about discrepancy review. Two authors but no editors answered the question about disliking discrepancy review. One author disliked that discrepancy review increased the overall word count and another disliked that the discrepancy review provided a recommendation to include information regarding missing data, but that this information was already reported in the manuscript. Across the questionnaires, no response indicated against conducting a trial of discrepancy review (e.g., unwillingness to use discrepancy review, excess time requirements, general dissatisfaction with the process).

### 3.6 Updated outcomes

Our preregistered outcome measure of discrepancies across each of the 18 dimensions was intended to answer two overarching questions: (1) whether authors address comments from discrepancy reviewers, and (2) whether a trial of discrepancy review could use this outcome measure. Based on information gained while performing this feasibility study, we revised our outcome measures to better answer these same questions.

#### 3.6.1 Procedural outcome

Our first revised outcome measure is whether published manuscripts addressed the specific recommendations in the discrepancy reports. For the first few discrepancy reports we prepared, each recommendation was directly linked to one of the 18 dimensions. However, we found this way of writing discrepancy reports cumbersome and switched to writing itemized comments that were not necessarily directly linked to one of the 18 dimensions. Thus, we assessed whether authors addressed the itemized recommendation of the discrepancy reports, but did not subdivide between the 18 dimensions

The discrepancy reports contained between 1-10 comments (mean = 4.8 SD = 2.7, median = 5, IQR = 3-7). Of the 59 comments in the 12 manuscripts that were published, 31 were fully addressed, 10 were partially addressed, 17 were not addressed, and 1 presented issues that the discrepancy reviewer later noticed did not need to be addressed. We categorized these 12 manuscripts into four bins: 6 addressed all or nearly all comments, 3 largely ignored discrepancy review and addressed very few comments (accounting for 13 of the 17 comments that were not addressed), 1 addressed some comments, and 2 were secondary publications associated with a clinical trial registration but did not explicitly state that some aspects of their study were not registered. One manuscript did not address any comments, likely due to an editorial misunderstanding: the ScholarOne portal requires that all reviewers provide a ‘Recommendation’, which we asked editors to ignore but which was considered in this one case. We coded several comments as partially addressed because the text in the published manuscript was imprecise. Inviting the discrepancy reviewers to review revised manuscripts may ensure that comments are more precisely addressed. In short, these data show that revised manuscripts often take into account the comments from discrepancy reviews. We do not plan to use this outcome measure for a trial. Rather we would recommend comparing endpoints between the control and experimental groups. Nonetheless, it provides evidence that discrepancy review has the potential to impact reporting quality.

#### 3.6.2 Potential trial outcome measures

We selected several other revised outcome measures that could feasibly be used in a trial. These include whether published manuscripts (1) are properly registered, (2) are transparently identified as secondary publications associated with a clinical trial registration, when relevant, (3) contain at least one primary outcome discrepancy, (4) contain at least one secondary outcome discrepancy, and (5) are assessed to have discrepancies that are important to address (see Table 2 for these results).

Inter-rater agreement varied greatly across these potential trial outcomes. Coders had perfect agreement (Cohen’s κ = 1.00) on whether there were issues in the registrations for submitted manuscripts (where 2/21 had issues) and had one disagreement (Cohen’s κ = 0.75) for published manuscripts (where 2/12 had issues). Coders agreed when identifying whether manuscripts were a secondary publication associated with a clinical trial registration for 18/21 submitted manuscripts (Cohen’s κ = 0.67) and 11/12 published manuscripts (Cohen’s κ = 0.8). Of the six published manuscripts linked with clinical trial registrations that were published, only two of them were the main report of the clinical trial. Coders agreed that neither had a primary outcome discrepancy and that both had at least one secondary outcome discrepancy. Coders’ subjective assessments of whether discrepancies were ‘quite important’, ‘somewhat important’, or ‘not important’ to address^8^ matched in 5 cases, were off by one category in 7 cases, and off by two categories in 1 case for submitted manuscripts (Cohen’s κ = 0.05). For published manuscripts, they matched in 4 cases, were off by one category in 1 case, and off by two categories in 2 cases (Cohen’s κ = -0.21). Inter-rater agreement was low for these subjective assessments, but differences in coding were easily resolved through discussion because one coder would often share information that the other coder had missed. In general, the coder who stated discrepancies were more important to address shifted the other coder’s rating. More precise instructions for these subjective assessments may improve inter-rater agreement to the point where it could be reliably used as an outcome measure in a trial.

## 4. Discussion

We found that (1) journals can incorporate discrepancy review into their manuscript handling procedures; (2) registrations were less precise and less comprehensive than our original discrepancy review process was designed for; (3) a semi-structured discrepancy review process was more feasible to implement than a rigidly-structured and comprehensive implementation; (4) authors addressed the majority of discrepancy reviewer comments, (5) clinical trial registrations and OSF registrations should be treated separately in a trial, as the former are generally more precise and the latter are generally more comprehensive; and (6) a trial of discrepancy review appears feasible in terms of the procedure and outcomes measures, particularly so for clinical trial registrations where primary and secondary outcome are consistently and clearly demarcated.

The outcome measures that could be used in a trial should depend on the types of registrations being reviewed. In contrast to OSF registrations, clinical trial registrations in our sample were always properly registered, but many manuscripts were secondary publications associated with a clinical trial registration. OSF registrations were generally less precise than clinical trials registrations, making it difficult to confidently claim an absence of primary or secondary outcome discrepancies. The lack of standardization in the content and precision of OSF registrations may also makes the process of awarding preregistered badges challenging. Box 2 outlines a potential design for a trial of discrepancy review.

### Box 2.

Potential design for a full trial on discrepancy review

#### Eligibility criteria

Manuscripts submitted to participating journals that include a clinical trial registration.

#### Design

Two-arm parallel-group randomized controlled trial. Randomized at the level of submitted manuscripts within each journal.

#### Intervention

Updated discrepancy review process.

#### Primary outcome

Absence of undisclosed primary outcome discrepancies. If a manuscript is a secondary publication associated with a clinical trial registration and is clearly labelled as such, it will be considered to have no undisclosed primary outcome discrepancies. This binary outcome will be coded from manuscripts accepted for publication.

#### Other outcomes

The time to prepare discrepancy reports. To be used to inform future cost-effectiveness analyses.

#### Sample size and analysis

This trial could be approached with a power analysis or precision analysis. For a power analysis, we would need to select a smallest effect size of interest. Determining this effect is difficult without knowing the cost that journals are willing to incur for discrepancy review and the cost of undisclosed discrepancies (e.g., in misdirected future research resources or reduced quality of patient care). A precision analysis, in contrast, remains agnostic regarding whether the effect is meaningful and could be used in follow-up cost-effectiveness analyses. Both power and precision analyses require assumptions about the proportion of manuscripts with discrepancies in both the control and experimental group. For the control group, we assume a base-rate of 33% of manuscripts with at least one primary outcome discrepancy, which is the point estimate from a relevant meta-analysis (TARG Meta-Research Group & Collaborators 2021). For the experimental group, we take a best guess that about 10% of publications will have at least one primary outcome discrepancy. To detect this difference between groups, a Fisher’s exact test would require 49 manuscripts per group (α = .05, power = .80). Alternatively, to estimate the difference between the two groups within a range of 20% (e.g., to show the experimental group has between 10-30% fewer manuscripts with primary outcome discrepancies), would require 120 manuscripts per group (95% confidence interval).^9^

Several questions remain unanswered. First, we have not established the degree to which general research knowledge and domain expertise—which the discrepancy reviewers did not have in relation to the manuscripts they reviewed—facilitate or improve discrepancy review. Second, our study was not designed to identify elements of discrepancy review that could be automated (e.g., with a standard email asking authors to ensure accurate reporting). Third, our study did not evaluate the feasibility of randomizing manuscripts to the discrepancy review intervention, which could pose a challenge when conducting a trial. Fourth, it remains unknown whether editors who received discrepancy reviews will become more aware of these issues and check for discrepancies in future manuscripts. Fifth, we did not assess how easy it would be to recruit discrepancy reviewers outside the context of a trial and whether journals are willing to incur those potential costs. Additionally, the respondents to our questionnaires were likely biased towards those who were more interested in discrepancy review.

Taken together, a trial of discrepancy review appears feasible if enough journals agree to participate. Such a trial may provide evidence for one potential intervention to improve reporting quality. At the same time, this intervention cannot directly improve study design or the quality of registrations and may add to the workload of researchers and reviewers. Parallel efforts that aim to improve quality early on in the research pipeline (e.g., pre-study peer review) could prove complementary to interventions, such as discrepancy review, that occur after study completion.

## Data Availability

The study protocol and materials were registered on 28 January 2021 at osf.io/5dh47/files. Discrepancies between this manuscript and the registered protocol are outlined in Supplementary Material A. Open data, codebooks, and the analysis script are temporarily stored at osf.io/25ebk. Upon acceptance for publication, these materials will be uploaded to the University of Bristol Research Data Repository. To maintain the confidentiality of the manuscripts we reviewed and the associated authors and editors, the discrepancy reviews will not be made publicly available.

## Supplementary material

www.osf.io/pe4wt

### Funding

Robert Thibault is supported by a general support grant awarded to METRICS from the Laura and John Arnold Foundation and a postdoctoral fellowship from the Fonds de recherche du Québec – Santé. Robbie Clark is supported by a SWDTP ESRC PhD studentship. Tom E. Hardwicke receives funding from the European Union’s Horizon 2020 research and innovation programme under the Marie Skłodowska-Curie grant agreement No. 841188. Jacqueline Thompson is supported by the UK Medical Research Council and previously by Jisc. Katie Drax is supported by the John Climax Benevolent Fund. Marcus Munafo, Robert Thibault, Jacqueline Thompson, Katie Drax, and Robbie Clark are all part of the MRC Integrative Epidemiology Unit (MC_UU_00011/7). The fellowship funders have no role in study design, data collection and analysis, decision to publish, or preparation of the manuscript.

### Contributions

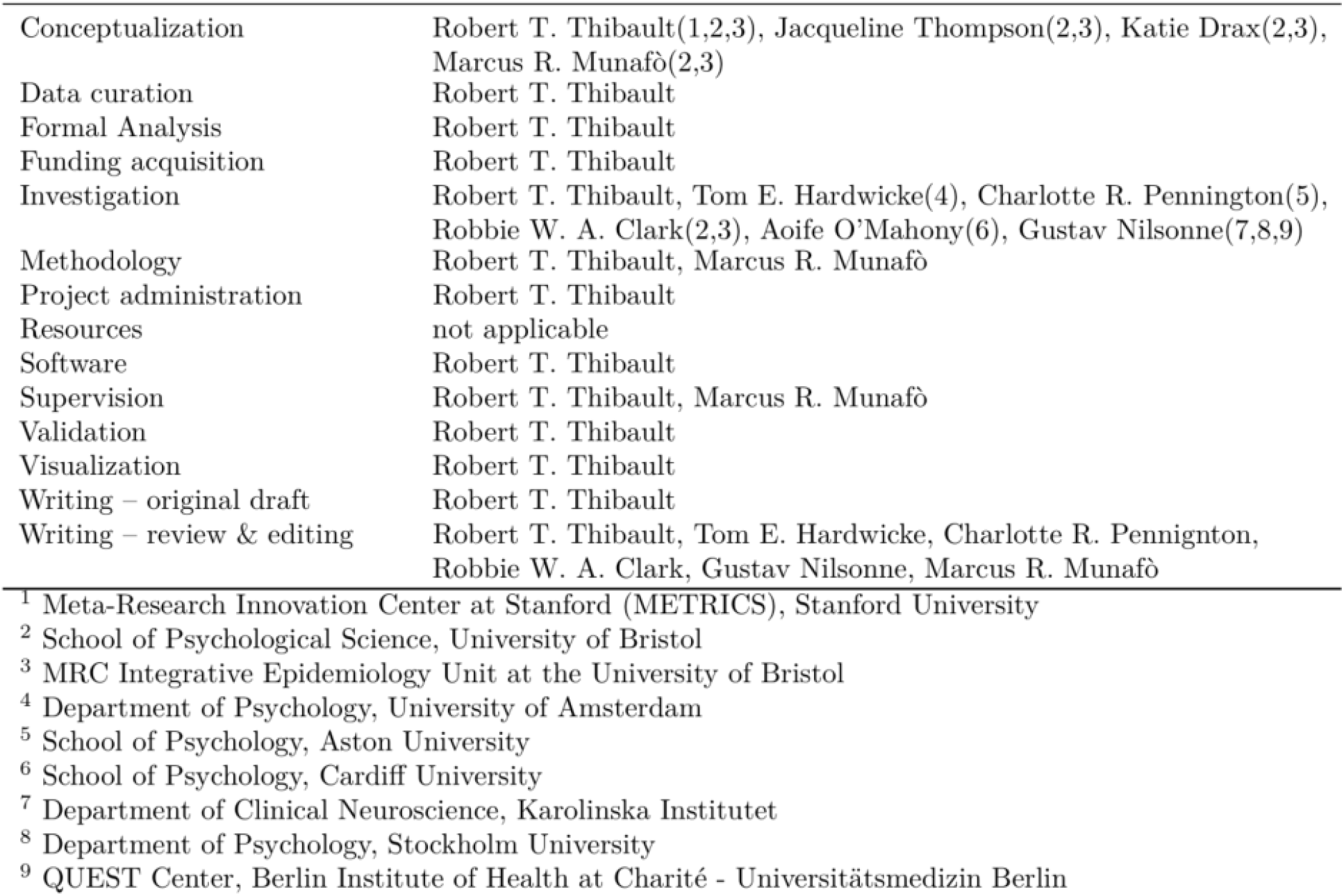

## Footnotes

1 The original version of the survey is available at osf.io/sfm2g. It was updated several times during the study and the most recent version is in Supplementary Material C.

2 The opt-in text reads: “[Journal name] is working with researchers to study and improve the peer review process and the quality of publications. This study is not intended to influence editorial decisions for accepting or rejecting a manuscript. Additional information will be provided for Associate Editors to consider but they will not view this until after they have made their initial editorial decision. If you agree to take part, your unblinded complete manuscript will be shared with the researchers, who will treat this information confidentially. You may drop out of the research study at any time without having to give a reason for doing so. If you would like your manuscript to contribute to this research, please check this box.”

3 Exact wording in Question 3.1 of Supplementary Material C

4 Operational definitions in Appendix A of Supplementary Material D

5 After we performed original discrepancy review on the first manuscript we received, we made changes to the extraction form. Thus, we exclude this manuscript from Table 1.

6 Clinical trials will often have a “main publication” that reports on the main questions a clinical trial is trying to answer. There will oftentimes be additional publications that use data from a clinical trial, but report analyses unrelated to the main purpose of the trial. In this manuscript we refer to these as “secondary publications associated with a clinical trial”.

7 This issue appears relatively common, perhaps because the interface is not particularly easy to understand.

8 See Supplementary Material F for operational definitions.

9 These calculations were made in R. The code is available in Supplementary Material G

## References

American Psychological Association, 2020. Preregistration [WWW Document]. https://www.apa.org. URL https://www.apa.org/pubs/journals/resources/preregistration (accessed 10.25.21).

Claesen, A., Gomes, S., Tuerlinckx, F., Wolf Vanpaemel, 2019. Preregistration: Comparing dream to reality. PsyArXiv. https://doi.org/10.31234/osf.io/d8wex

COS, 2021. Select a Registration Template [WWW Document]. OSF Guides. URL https://help.osf.io/hc/en-us/articles/360019738794-Select-a-Registration-Template (accessed 10.25.21).

COS, 2019. Badges to Acknowledge Open Practices [WWW Document]. URL https://osf.io/tvyxz/wiki/1.%20View%20the%20Badges/ (accessed 10.25.21).

COS, 2016. Badges to Acknowledge Open Practices [WWW Document]. URL https://osf.io/tvyxz/wiki/2.%20Awarding%20Badges/ (accessed 10.25.21).

Goldacre, B., Drysdale, H., Dale, A., Milosevic, I., Slade, E., Hartley, P., Marston, C., Powell-Smith, A., Heneghan, C., Mahtani, K.R., 2019. COMPare: A prospective cohort study correcting and monitoring 58 misreported trials in real time. Trials 20, 1–16. https://doi.org/10.1186/s13063-019-3173-2

Hardwicke, T.E., Wagenmakers, E.-J., 2021. Preregistration: A pragmatic tool to reduce bias and calibrate confidence in scientific research. https://doi.org/10.31222/osf.io/d7bcu

ICMJE, 2019. Recommendations for the Conduct, Reporting, Editing, and Publication of Scholarly Work in Medical Journals.

Jones, C.W., Adams, A., Weaver, M.A., Schroter, S., Misemer, B.S., Schriger, D., Platts-Mills, T.F., 2019. Peer reviewed evaluation of registered end-points of randomised trials (the PRE-REPORT study): Protocol for a stepped-wedge, cluster-randomised trial. BMJ Open 9, 1–8. https://doi.org/10.1136/bmjopen-2018-028694

Lakens, D., 2019. The Value of Preregistration for Psychological Science: A Conceptual Analysis (preprint). PsyArXiv. https://doi.org/10.31234/osf.io/jbh4w

Lash, T.L., Vandenbroucke, J.P., 2012. Should Preregistration of Epidemiologic Study Protocols Become Compulsory? Reflections and a Counterproposal. EPIDEMIOLOGY 23, 184–188. https://doi.org/10.1097/EDE.0b013e318245c05b

Mathieu, S., Chan, A.W., Ravaud, P., 2013. Use of Trial Register Information during the Peer Review Process. PLoS ONE 8, 2–5. https://doi.org/10.1371/journal.pone.0059910

NC3Rs, 2021. NC3Rs Funding Schemes Applicant and Grant Holder Handbook.

Nosek, B.A., Ebersole, C.R., DeHaven, A.C., Mellor, D.T., 2018. The preregistration revolution. PROCEEDINGS OF THE NATIONAL ACADEMY OF SCIENCES OF THE UNITED STATES OF AMERICA 115, 2600–2606. https://doi.org/10.1073/pnas.1708274114

TARG Meta-Research Group & Collaborators, 2021. Estimating the prevalence of discrepancies between study registrations and publications: A systematic review and meta-analyses (preprint). Health Systems and Quality Improvement. https://doi.org/10.1101/2021.07.07.21259868

van den Akker, O., 2019. The effectiveness of preregistration: Assessing preregistration strictness and preregistration-paper consistency. OSF.

World Health Organization, n.d. WHO Trial Registration Data Set (Version 1.3.1) [WWW Document]. URL https://www.who.int/clinical-trials-registry-platform/network/who-data-set (accessed 10.25.21).

World Medical Association, 2013. Declaration of Helsinki – Ethical Principles for Medical Research Involving Human Subjects.

Zarin, D.A., Tse, T., Williams, R.J., Rajakannan, T., 2017. Update on Trial Registration 11 Years after the ICMJE Policy Was Established. New England Journal of Medicine 376, 383–391. https://doi.org/10.1056/NEJMsr1601330

