## Supplementary Material for "Discrepancy review: A feasibility study of a novel peer review intervention to reduce undisclosed discrepancies between registrations and publications"

**Supplementary Material A.** Protocol amendments.

**Supplementary Material B.** Additional information on the original discrepancy review process.

**Supplementary Material C.** Qualtrics form guiding the original discrepancy review process.

**Supplementary Material D.** Instructions for the original discrepancy review process, including our operational definitions of major, minor, and negligible issues.

**Supplementary Material E.** Instructions guiding the updated discrepancy review process.

**Supplementary Material F.** Qualtrics form of the follow-up questions after discrepancy review, including operational definitions of the importance of addressing discrepancies.

**Supplementary Material G.** Power analysis and precision analysis code.

**Supplementary Material A.** Protocol amendments.

The study protocol was pre-registered on 28 January 2021 (osf.io/5dh47). We conducted a feasibility study where we iteratively refined our intervention based on insights gained during data collection, and with the aim of developing an intervention that could later be used in a trial. Thus, making amendments to our protocol was part of the study objectives. Supplementary Table A1 outlines the amendments between our registered protocol and final report.

| **Supplementary Table A1**. Protocol amendments. | | | | |
| --- | --- | --- | --- | --- |
| **Issue** | **Preregistered protocol** | **Amendment** | **Rationale** | **Was the decision made after observing relevant study results?** |
| Dimensions assessed in the original discrepancy review process | We outlined 18 dimensions. | We stopped coding for whether statistical software was reported and started coding for secondary outcomes. In the protocol we call these “elements” rather than “dimensions”. | Coding for statistical software seemed too critical and we felt the need to separately code for secondary outcomes, particularly for clinical trial registrations. | Yes, it was made after performing a few discrepancy reviews. |
| Coding form | The original form we used to help prepare a discrepancy report is available at osf.io/sfm2g/ | We updated this form several times in order to refine discrepancy review. The final version of this form is in Supplementary Material C. | This was one of our preregistered study objectives. | Yes. We did this throughout data collection. |
| Undisclosed discrepancies | The protocol did not mention we were looking exclusively for undisclosed discrepancies. | The instruction sheets for discrepancy reviewers asked them to look only at undisclosed discrepancies. | This information was intended to be included in the protocol. That it is not included was an oversight. | Yes, when preparing the instruction sheet for discrepancy reviewers. Although this information was already known by our team and the discrepancy reviewers. |
| Instructions to discrepancy reviewers | The protocol did not include specific instructions to reviewers, beyond what is in the protocol appendices. | We prepared a specific document to walk discrepancy reviewers through the process of conducting discrepancy review. Available in Supplementary Material D & E. | This seemed like the most efficient and consistent way of training discrepancy reviewers. | Yes. The project lead (RTT) wrote this after reviewing a few submitted manuscripts himself. |
| Updated discrepancy review | The protocol only outlined the original discrepancy review process. | We developed a more streamlined discrepancy review process that differed substantially in the time taken and depth of information collected. Available in Supplementary Material E. | The original discrepancy review process seemed impractical to implement and run a trial on. Refining discrepancy review was one of our registered study objectives. | Yes. This was developed after a first coder reviewed 16 submitted manuscripts and a second coder reviewed 8 submitted manuscripts. |
| Discrepancy reports | The protocol contained a systematic method for producing discrepancy reports. Template available at osf.io/786eb/ | Discrepancy reviewers began to prepare shorter itemized reports that were less structured and could report on any issue. | The original discrepancy reports seemed too cumbersome for authors and editors to digest quickly. Refining discrepancy review was one of our registered study objectives. | Yes, after reviewing several submitted manuscripts. |
| Potential trial outcome measures | The protocol said we would test whether the 18 dimensions were reasonable trial outcomes. | We tested several additional outcomes that might be possible to use in a trial. | The original outcomes seemed unfit for use in a trial. | Yes, after reviewing most of the submitted manuscripts. |
| Prospective registration and preregistration | We use the term preregistration broadly in the protocol | In the manuscript, we largely use the term registration and distinguish between OSF registrations and clinical trial registrations. | Given the difference we found between registrations on the OSF and clinical trials registrations, we felt the need to distinguish them in our writing. Clinical trials don’t often use the term preregistration. | Yes, when writing the manuscript. |
| Reviewer questionnaire to be completed after performing discrepancy review | “Discrepancy reviewers also complete a short questionnaire immediately after completing the discrepancy review” | We modified this questionnaire several times. The data from these questionnaires are included in the Table 2 data file, rather than a separate data file for the questionnaire alone. | We added questions about the importance of addressing discrepancies, to explore potential outcome measures for a trial. | Yes. We modified the questionnaire a few times during data collection. |
| Reviewer questionnaire to be completed after a manuscript is accepted or rejected | The protocol states: “After a manuscript that underwent discrepancy review is rejected or accepted for publication, action editors, manuscript authors, and discrepancy reviewers will each receive a feedback questionnaire to complete.” | Reviewers only completed a questionnaire immediately upon completing discrepancy review. However, much of the information reviewers would have been asked at this stage was included in our manuscript after a group discussion with all the discrepancy reviewers. | The manuscript submission system from the journals we partnered with did not inform us when a final editorial decision was made. | This was discovered when beginning to prepare the manuscript. |
| Timing of author questionnaire | “After a manuscript that underwent discrepancy review is rejected or accepted for publication, action editors, manuscript authors, and discrepancy reviewers will each receive a feedback questionnaire to complete.” | Initial editorial decisions included text that informed authors about our study and asked them to complete the questionnaire. | It was easier to inform the author about the study and ask for their feedback at the same time. | No. It was made before reviewing any submitted manuscripts. |
| Number of registered manuscripts submitted | Objective 1a: “Document the number of preregistered manuscripts submitted to participating journals.” | Only two journals provided us with this information. | We asked journals for this information, but some could not provide it. | NA |
| Assigning discrepancy review | Objective 3f: “Establish and improve the process of assigning discrepancy reviewers to manuscripts.” | We assigned discrepancy reviews based on reviewer availability. | We had a limited number of discrepancy reviewers with limited availability. | Decided while reviewing submitted manuscripts. |
| Contributorship | Projected contributorship roles are outlined. | Additional contributors joined the team. | This was planned “Additional contributors will likely be recruited to perform the discrepancy review.” | While reviewing submitted manuscripts. |

**Supplementary Material B.** Additional information on the original discrepancy review process.

**Supplementary Table B1.** Shortcomings in the original discrepancy review process and how we addressed them. The content of this table stems from feedback received from the five discrepancy reviewers involved in this project.

| **Issue with original method** | **Solution implemented for updated discrepancy review process** |
| --- | --- |
| Discrepancy review takes too long. | We used the original discrepancy review process for two purposes: to act as the intervention and to evaluate outcomes. Whereas the outcome evaluation stage needs high precision, some errors are acceptable in the intervention stage—authors can rebut anything they disagree with throughout the peer review process. Thus, we developed a more streamlined version of discrepancy review that aims to maximize benefits in relation to the time required to perform the review. |
| Some manuscripts report only non-registered outcome measures (e.g., an exploratory analysis of a clinical trial where the main registered outcomes were already published elsewhere). Discrepancy review seems inappropriate for these manuscripts. | We added a triage process to identify these manuscripts. Discrepancy reviewers asked authors of these manuscripts to explicitly state that the outcomes and analyses in these manuscript were not registered and represent explorations of data collected from a registered trial that was previously published elsewhere. |
| We coded dimensions categorically as (i) not reported, (ii) reported but non-reproducibly, or (iii) reported reproducibly. This coding is highly subjective and fails to capture important differences between reporting that is poor versus reporting that is very good but not quite reproducible. | We removed the requirement to code whether dimensions are reported reproducibly. Discrepancy reviewers may still provide comments regarding poor reporting if they wish to. |
| The process can feel pedantic. Many discrepancies may have only a negligible or minor impact on reporting quality. This could negatively impact how authors and editors engage with discrepancy review. | Our updated intervention focuses on dimensions where discrepancies are more likely to have substantial impact including hypotheses, outcome measures, and analyses. Discrepancy reviewers may still comment on other dimensions if they choose to. |
| Some dimensions are not applicable to many manuscripts (e.g., randomization in an observational study) and few registrations contain details on many of the dimensions (e.g., violations of statistical assumptions) | Our updated intervention focuses on dimensions where discrepancies are more likely to have substantial impact including hypotheses, outcome measures, and analyses. Discrepancy reviewers may still comment on other dimensions if they choose to. |
| Many discrepancy reviewer recommendations are similar across manuscripts and seem to fill in gaps in manuscript authors’ knowledge regarding the importance of accurate reporting in relation to registrations. | To save time, we drafted a few standard comments that can be copy-pasted into discrepancy reports. This remark also raises the idea that instructions sent to the authors *before* submission could lead to submitted manuscripts with fewer discrepancies. Such an intervention could be tested alone or in parallel with discrepancy review. |

**Supplementary Table B2.** Reporting and discrepancies across the 18 dimensions included in the original discrepancy review process (second coder, n = 6). Reporting was coded as 0 (not reported), 1 (reported, but unclear), or 2 (reported clearly). This table presents data from the second coder for the 6 manuscripts that underwent the original discrepancy review process and were not a secondary publication associated with a clinical trial registration.

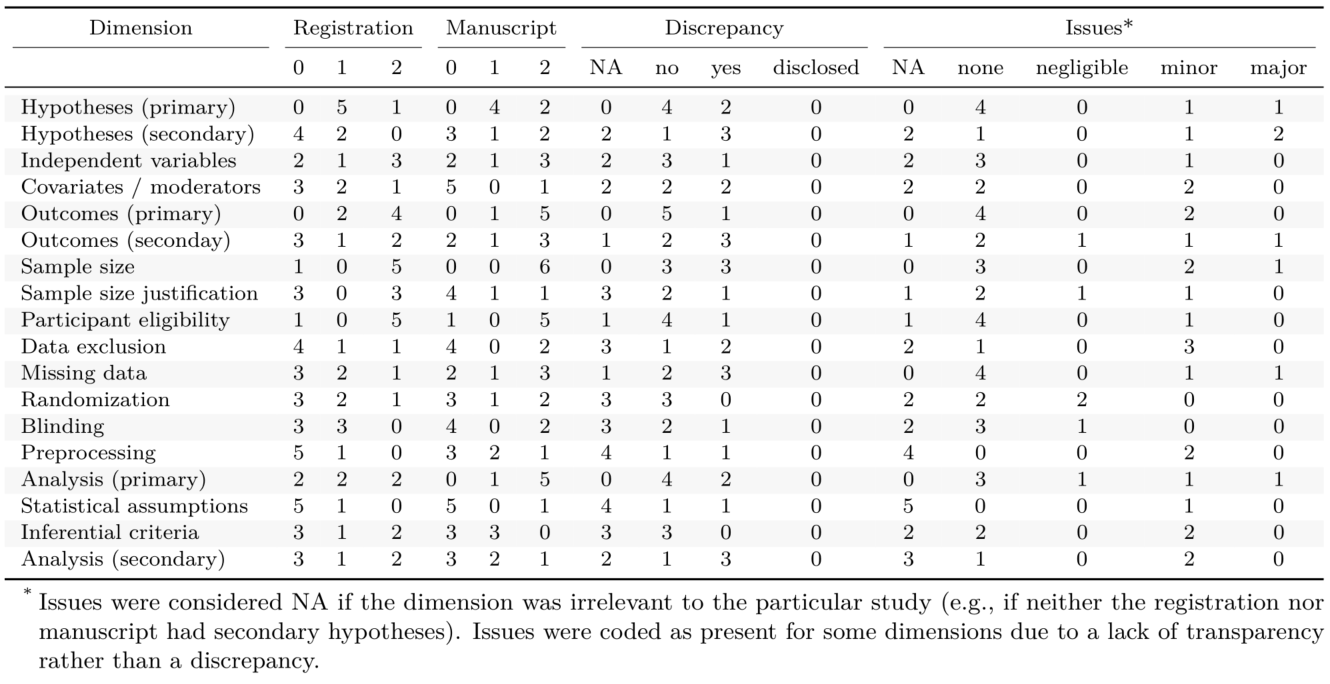

**Supplementary Table B3.** Inter-rater agreement across the 18 dimensions included in the original discrepancy review process (n = 6). This table lists the number of manuscripts where the first and second reviewers selected the same option (6 is perfect agreement). For items in the first four columns, reviewers had three options to select from. For the item in the final column, reviewers had five options.

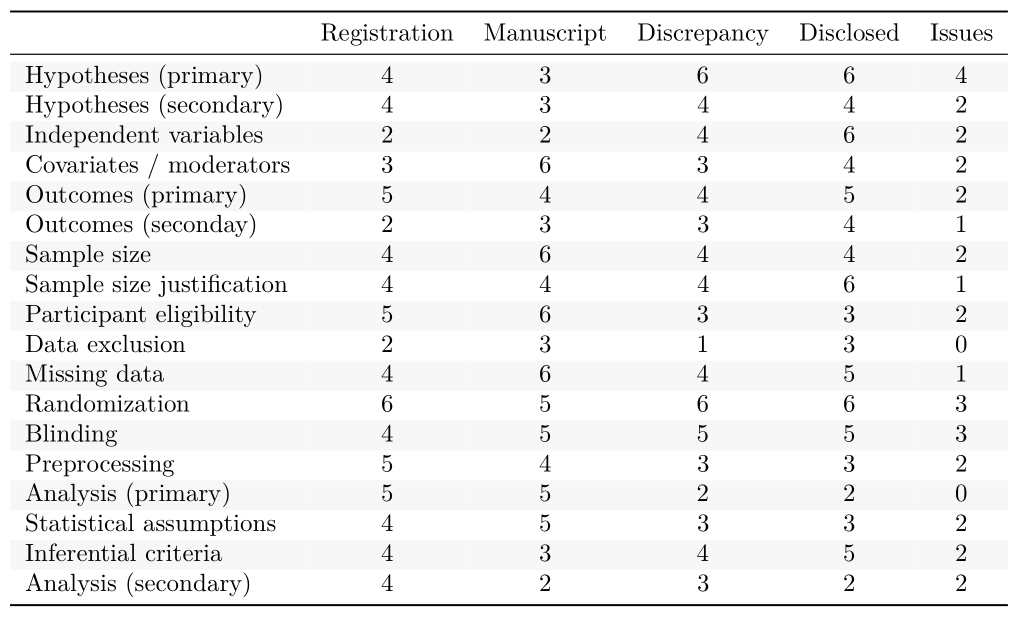

**Supplementary Material C.** Qualtrics form guiding the original discrepancy review process.

Many of the terms we use in this feasibility study are operationalized within this Qualtrics form.

**Standardized discrepancy form – 210324**

**Survey Flow**

**Block: Check (19 Questions)**

**Branch: New Branch**

**If**

**Invalid Logic Click Here to Edit Logic**

**Branch: New Branch**

**If**

**Invalid Logic Click Here to Edit Logic**

**Standard: Hypotheses / Aims (7 Questions)**

**Standard: Variables (10 Questions)**

**Standard: Analyses (16 Questions)**

**Standard: Model (9 Questions)**

**Standard: Prereg badge (2 Questions)**

**Standard: Additional comments (1 Question)**

| Page Break |
| --- |

**Start of Block: Check**

Q1.1 The design of this questionnaire was adapted from the work of Olmo van den Akker (https://osf.io/ta3yd/) and Bakker et al. (https://psyarxiv.com/cdgyh/). Some of the text is copied directly from these sources. Some of the text is adapted.

Q1.2 Enter the **URL where the preregistration is stored.** 
This is to facilitate matching between the forms for the preregistration and the submitted manuscript.

________________________________________________________________

Q1.3 Which document are you reviewing?

- preregistration (1)
- submitted manuscript (2)

*Skip To: Q1.19 If Which document are you reviewing? = submitted manuscript*

Q1.4 Do you have **a copy of both the preregistration and the submitted manuscript**. 
For the purpose of this form, we define a preregistration as a publicly available document that includes elements of a study protocol and that was archived before the beginning of a study. This definition includes pre-analysis plans and prospective registrations.

- yes (1)
- no (2)

*Display This Question:*

*If Do you have a copy of both the preregistration and the submitted manuscript.  For the purpose of... = no*

Q1.5 Restart this questionnaire once you have a copy of both the preregistration and the submitted manuscript.

*Skip To: End of Survey If Restart this questionnaire once you have a copy of both the preregistration and the submitted man... Is Displayed*

Q1.6 Read the abstract of the manuscript and quickly scan the preregistration. 
**Are the manuscript and preregistration sufficiently similar** that you would consider that they reflect the same study?
 
If you deem them to be "different studies" than you do NOT need to perform discrepancy review.
 
We do not yet have a strict operationalization of what qualifies as "different studies". Here is some information to help you decide:
 
(1) If the manuscript points to a registration, but the only thing that overlaps between the manuscript and the registration is the participant sample, then we consider these to be different studies. For example, a manuscript may report only on qualitative or demographic data that was collected during a clinical trial, but no info about the qualitative or demographic data appear in the clinical trial registry (i.e., not in the aims or the outcomes of the trial registration).
 
(2) Do you think it would be more appropriate for the manuscript to include:
(i) an overarching statement along the lines of "This manuscript reports a secondary/exploratory analysis of a study registered at [registration ID], but none of the outcome measures or analysis in this manuscript are included in the registration and none of the registered outcome measures or analyses are included in the manuscript"; OR
(ii) a disclosure for every discrepancy between the registration and manuscript (this could be in the manuscript text, or in an appendix that outlines all the discrepancies).
If you think (i) is more appropriate, than we consider these to be "different studies" and you should select "No" from the options below.

- Yes (1)
- No (2)

*Display This Question:*

*If Read the abstract of the manuscript and quickly scan the preregistration.  Are the manuscript and... = No*

Q1.7 Because you have deemed the preregistration and manuscript to be reflect "different studies", you do not need to perform discrepancy review. Instead, write a few sentences that you would suggest the authors include in their manuscript. Here is an example comment:

*This study presents a further exploration of the same sample used in a previous publication, but none of the outcomes or analyses in the submitted manuscript are registered. I suggest that the manuscript includes a statement along the lines of: “The inclusion and exclusion criteria for the sample are registered at [insert link]. The intervention conditions, outcome measures, and analyses reported in this manuscript are not registered, and none of the registered outcomes are reported in the manuscript.”*

*Skip To: End of Survey If Because you have deemed the preregistration and manuscript to be reflect "different studies", you... Is Displayed*

Q1.8 Are there **multiple versions of the preregistration** on the web page where you are accessing the preregistration?
If yes, use Version 1 throughout this form (because we want to compare the manuscript to the prospectively registered information, rather than to retrospectively registered information). If it is clear that a later version of the registration includes updates that were made before the study began, use that version. For example, Verison 1 of a registration may only include basic information and then a few months later Version 2 may be uploaded and include the outcome measures and analysis plan, but the study has yet to begin. At the moment, we have not yet strictly operationalized how this should be done. Use your judgement and feel free to email Robert if you're uncertain. You may choose to ask the authors to state in their manuscript at what stage the study was at when certain content was uploaded to the registration.
 
If the registration is on clinicaltrials.gov, use *control-f* and search for "History of changes". You can then merge and compare different versions of the registration.

- yes (1)
- no (2)

Q1.9 Enter the **upload date** of the version of the preregistration you are reviewing.

 If only the month and year are provided, input "00" as the date (yyyy-mm-dd).

________________________________________________________________

Q1.10 **Is the preregistration date questionably close to the submission date?**
Such that information in the preregistration may have been documented after the study began? The date range that could be considered questionable will vary depending on the type of study (e.g., a data collection period of two years versus an online survey of 100 participants).

- yes (1)
- unsure (2)
- no (4)

Q1.11
Identify the **template or format of the registration** (e.g., unrestricted, clinicaltrials.gov, OSF challenge).

________________________________________________________________

Q1.12 Where is the preregistration stored?

- ClinicalTrials.gov (5)
- OSF Registries (the words "OSF Registries" will be in the top left corner of the web page) (1)
- OSF Home (the words "OSF Home" will be in the top left corner of the web page). Note, this fails to qualify as a registration because the file is not permanent. In this case, make a recommendation to the authors that they make a permanent version of their preregistration using the OSF REGISTRIES website. You should still perform discrepancy review on this document. (2)
- AsPredicted (3)
- PROSPERO (4)
- EUCRT (6)
- AEA RCT (7)
- RIDIE (8)
- WHO International Clinical Trials Registry Platform (9)
- other (10) ________________________________________________

*Skip To: End of Block If Where is the preregistration stored? != other*

Q1.13 **Permanent**: The preregistration cannot be removed.

- true (1)
- unsure (2)
- false (4)

Q1.14 **Read-only**: The preregistration cannot be edited.

- true (1)
- unsure (2)
- false (4)

Q1.15 **Time-Stamp**: There is a time stamp of the day the preregistration is created (uploaded).

- true (1)
- false (2)

Q1.16 **Public**: The preregistration is accessible to anyone with an internet connection (i.e., you can access it for free and when not connected to your institutions internet).

- true (1)
- unsure (2)
- false (3)

Q1.17 **Non-ambiguously accessible**: The amount of time and effort to find the preregistration, or to reconstruct all information to one preregistration, is minimal (i.e., it took a maximum of 5 minutes to access the preregistration and, if it was divided over multiple documents, to assemble all relevant information).

- true (1)
- false (2)

Q1.18 **In third party repository**: The preregistration is stored on an third party repository (e.g., not the authors personal website).

- true (1)
- false (2)

*Skip To: End of Block If In third party repository: The preregistration is stored on an third party repository (e.g., not... , true Is Displayed*

Q1.19 Does the submitted **manuscript contain a link that clearly directs the reader to the preregistration** OR contain a non-ambiguous Registration ID (e.g., for clinicaltrials.gov or PROSPERO) that can be easily used to find the registration?

- Yes, the study preregistration is mentioned and accessible through a unique and persistent identifier (i.e., a URL link leads to a single file which is readily identifiable as the preregistration and there is little risk that the link will be dead in the future) OR unique and persistent Registration ID. (1)
- No, the study preregistration is mentioned, but is not accessible through a unique identifier (the link leads to multiple files and it is unclear which ones are relevant) or persistent identifier (there is risk that the link will be dead in the future), or you could eventually find the preregistration but it took at least a few minutes or expert knowledge (e.g., high familiarity with the osf.io infrastructure). (2)
- No, the study preregistration is not mentioned (3)

**End of Block: Check**

**Start of Block: Hypotheses / Aims**

Q2.1 Throughout this form you will be asked to copy-paste text from the document you are reviewing. In some instances, a single sentence may be sufficient. In other instances, you may need to copy multiple paragraphs. Copy as much text is necessary to try to save yourself from needing to reopen the preregistration or manuscript. Nonetheless, it's often still necessary to reopen the preregistration or manuscript to double check things.
If you would like to add notes to the text you copy, place your thoughts in [square brackets]. These may help when it comes time to compare the excerpts from the preregistration and submitted manuscript.
---------------------------------------------------------

Q2.2 Please identify all hypotheses in the document. If there are no "hypotheses", identify the "aims" or "objectives".  Please include both hypotheses and aims where the variables are presented on a conceptual level, as well as hypotheses and aims where the variables are presented using a specific operationalization. If a single hypothesis is explained on both a conceptualized and operationalized level (and it is clear that they are the same hypothesis) choose the operationalized hypothesis. If it is unclear if they are the same hypotheses use the information from both the conceptual and operationalized levels.  The hypothesis or aim may be phrased using the terms "expect”, “predict”, “anticipate”, “theor*”, “hypothes*”, "aim", "examine", "investigate", "answer", "objective", "resesarch question" or other terms.
 **Determine whether the document distinguishes between primary/main/prioritized versus secondary/other/additional/non-prioritized hypotheses.** A numbering of hypotheses alone doesn't necessarily indicate primary versus secondary hypotheses. Note, many registrations do not distinguish between primary and secondary hypotheses. There may be multiple primary hypotheses and multiple secondary hypotheses.

Q2.3 Is at least one hypothesis or aim specified?

- Yes, only one is made (1)
- Yes, multiple are made with a clear priority (e.g., primary vs. secondary hypotheses) (2)
- Yes, multiple are made and the priority can be reasonably assumed (3)
- Yes, multiple are made and the priority CANNOT be reasonably assumed (4)
- No hypothesis or aim is specified (5)

*Skip To: End of Block If Is at least one hypothesis or aim specified? = No hypothesis or aim is specified*

Q2.5 Is the direction of the **primary hypothesis(es) / aim(s)** specified?
 Note, for the purposes of this form, if there is no prioritization of hypotheses, we will consider all hypotheses as "primary". Only select "yes", if the direction of **all** primary hypotheses / aims are specified.

- yes (1)
- no (2)

Q2.6 Copy-paste the main or **primary hypothesis(es) / aim(s**) in the text box below.

________________________________________________________________

________________________________________________________________

________________________________________________________________

________________________________________________________________

________________________________________________________________

*Display This Question:*

*If Is at least one hypothesis or aim specified? = Yes, multiple are made with a clear priority (e.g., primary vs. secondary hypotheses)*

*Or Is at least one hypothesis or aim specified? = Yes, multiple are made and the priority can be reasonably assumed*

Q2.9 Is the direction of the secondary hypothesis(es) / aim(s) specified?
If the direction of at least one secondary hypothesis is not indicated, select "no".

- yes (1)
- no (2)

*Display This Question:*

*If Is at least one hypothesis or aim specified? = Yes, multiple are made with a clear priority (e.g., primary vs. secondary hypotheses)*

*Or Is at least one hypothesis or aim specified? = Yes, multiple are made and the priority can be reasonably assumed*

Q2.10 Copy-paste the **secondary** hypothesis(es) / aim(s) in the text box below.

________________________________________________________________

________________________________________________________________

________________________________________________________________

________________________________________________________________

________________________________________________________________

**End of Block: Hypotheses / Aims**

**Start of Block: Variables**

Q3.1 **A note on terminology.**
 This form asks if several items in the preregistration and manuscript are described **sufficiently**. In other words, we are asking if the document provides enough detail that you think you could perform this part of the study yourself. Feel free to interpret the term "sufficiently" as "reproducible".  To be sufficient, the item should be described including all steps that will be taken (it should be specific), and each of the described steps should allow only one interpretation or implementation (it should be precise). If the item is described in a manner that is not fully specific and precise, but in a manner that is clear enough for you to conduct the study, consider it "sufficient". In some instances, you will not have the domain expertise to be confident about whether the description is sufficient. In these cases, take your best guess at whether you think the information provided would be sufficient for another expert in that domain to run the study.

Q3.2 Does the document describe any **independent variable(s), exposure variable(s), or study/experimental grouping** (these can be manipulated or non-manipulated)?
Are they described sufficiently for you to run the study?

- Yes, sufficiently (1)
- Yes, **IN**sufficiently (2)
- No, and these variables are, or are likely to be, relevant (5)
- No, and these variables are, or are likely to be, irrelevant (NA) (e.g., there is only one group of participants). (6)

Q3.3 Copy-paste the text describing the **independent variable(s), exposure variable(s), or study/experimental grouping** in the text box below.
 Enter "relevant" or "NA" if you chose either of the "No" options to the previous question.

________________________________________________________________

________________________________________________________________

________________________________________________________________

________________________________________________________________

________________________________________________________________

Q3.4 Does the document describe any **control variable(s), covariates, confounders, or moderators**?
 For the purposes of this study, we do not consider demographic characteristics (e.g., the details often included in "Table 1": age, gender, etc) to be control variable(s), covariates, confounders, or moderators for this question unless the document explicitly states how this demographic information will be used as such.

- Yes, sufficiently (1)
- Yes, **IN**sufficiently (2)
- No, and these variables are, or are likely to be, relevant (3)
- No, and these variables are, or are likely to be, irrelevant (NA) (e.g., there is only one group of participants). (4)

Q3.5 Copy-paste the text describing the **control variable(s), covariates, confounders, or moderators** in the text box below.
 Enter "relevant" or "NA" if you chose either of the "No" options to the previous question.

________________________________________________________________

________________________________________________________________

________________________________________________________________

________________________________________________________________

________________________________________________________________

Q3.6 Does the document specify **outcome measures**, dependent variables, or endpoint measures?
Determine whether the document distinguishes between primary and secondary outcomes. Note, many registrations do not make this distinction. 
For the manuscript, extract the outcomes from the Results section rather than the Methods section. We advise this because sometime a manuscript will mention outcomes in the methods section, but never reported them in the results section.

 

- Yes, only one is specified (1)
- Yes, multiple are specified with a clear priority (2)
- Yes, multiple are specified and the priority can be reasonably assumed (3)
- Yes, multiple are specified and the priority CANNOT be reasonably assumed (4)
- No outcome measures are specified (5)

*Skip To: End of Block If Does the document specify outcome measures, dependent variables, or endpoint measures? Determine... = No outcome measures are specified*

Q3.7
 This form considers all outcomes to be **primary outcomes** unless they are clearly demarcated as, or can reasonably be assumed to be, "secondary", "tertiary", "other", "additional", or are non-prioritized in another way. Does the text specify how this (or these) **primary outcome(s)** will be measured?  If there are multiple primary outcomes, and at least one of them is insufficiently described, select "Yes, INsufficiently"

- Yes, sufficiently (1)
- Yes, **IN**sufficiently (2)
- No (3)

Q3.8 Copy-paste the text describing the **primary outcome measure(s)** in the text box below.
 If there are multiple primary outcome measures, copy-paste the text for all of them.

________________________________________________________________

________________________________________________________________

________________________________________________________________

________________________________________________________________

________________________________________________________________

*Display This Question:*

*If Does the document specify outcome measures, dependent variables, or endpoint measures? Determine... = Yes, multiple are specified with a clear priority*

*Or Does the document specify outcome measures, dependent variables, or endpoint measures? Determine... = Yes, multiple are specified and the priority can be reasonably assumed*

Q3.9 This form considers outcomes to be **secondary outcomes** only if they are clearly demarcated as, or can reasonably be assumed to be, "secondary", "tertiary", "other", "additional", or are non-prioritized in another way.
Does the text specify how the **secondary outcome(s)** will be measured? 

If there are multiple secondary outcomes, and at least one of them is insufficiently described, select "Yes, **IN**sufficiently"

- Yes, sufficiently (1)
- Yes, **IN**sufficiently (2)
- No (3)

*Display This Question:*

*If Does the document specify outcome measures, dependent variables, or endpoint measures? Determine... = Yes, multiple are specified with a clear priority*

*Or Does the document specify outcome measures, dependent variables, or endpoint measures? Determine... = Yes, multiple are specified and the priority can be reasonably assumed*

Q3.10 Copy-paste the text describing the **secondary outcome measure(s)** in the text box below.
 If there are multiple secondary outcome measures, copy-paste the text for all of them.

________________________________________________________________

________________________________________________________________

________________________________________________________________

________________________________________________________________

________________________________________________________________

**End of Block: Variables**

**Start of Block: Analyses**

Q4.1 Does the document state the **sample size**?

- Yes, a specific number or range (1)
- Yes, a minimum or maximum (2)
- No (3)

Q4.2 Copy-paste the text describing the **sample size** in the text box below. 
Type "NA" if not applicable.

________________________________________________________________

________________________________________________________________

________________________________________________________________

________________________________________________________________

________________________________________________________________

Q4.3 Does the document provide a **justification for their sample size**.
 For example, a power analysis, Bayesian sequential sampling calclaion, sample size determination, reference to a previous study, or any other explicit statement about why they chose this sample size.
Is it sufficiently describe to be reproducible?

- Yes, sufficiently (1)
- Yes, **IN**sufficiently (2)
- No (3)

Q4.4 Copy-paste the text **justifying the sample size** in the text box below.
 Type "NA" if not applicable.

________________________________________________________________

________________________________________________________________

________________________________________________________________

________________________________________________________________

________________________________________________________________

Q4.5 Does the document describe **inclusion or exclusion criteria** that will be used to select **participants**?

- Yes, sufficiently (1)
- Yes, **IN**sufficiently (2)
- No, and inclusion or exclusion criteria for participants are, or are likely to be, relevant (3)
- No, and inclusion or exclusion criteria for participants are, or are likely to be, **NOT** relevant (NA) (4)

Q4.6 Copy-paste the text describing the **participant inclusion or exclusion criteria** in the text box below.
 Enter "relevant" or "NA" if you chose either of the "No" options to the previous question.

________________________________________________________________

________________________________________________________________

________________________________________________________________

________________________________________________________________

________________________________________________________________

Q4.7 Does the document describe **inclusion or exclusion criteria** that will be used to select which collected **data** are used in the analysis (this question includes references to "outliers")?

- Yes, sufficiently (1)
- Yes, **IN**sufficiently (2)
- No, and inclusion or exclusion criteria for data are, or are likely to be, relevant (3)
- No, and inclusion or exclusion criteria for data are, or are likely to be, **NOT** relevant (NA) (4)

Q4.8 Copy-paste the text describing the data inclusion or exclusion criteria in the text box below.
Enter "relevant" or "NA" if you chose either of the "No" options to the previous question.

________________________________________________________________

________________________________________________________________

________________________________________________________________

________________________________________________________________

________________________________________________________________

Q4.9 Does the document describe how the study deals with **incomplete or missing data**?

- Yes, sufficiently (1)
- Yes, **IN**sufficiently (2)
- No, and incomplete or missing data are, or are likely to be, relevant (3)
- No, and incomplete or missing data are, or are likely to be, **NOT** relevant (NA) (4)

Q4.10 Copy-paste the text describing **incomplete or missing data** in the text box below.
 Enter "relevant" or "NA" if you chose either of the "No" options to the previous question.

________________________________________________________________

________________________________________________________________

________________________________________________________________

________________________________________________________________

________________________________________________________________

Q4.11 Does the document describe how **randomization** of the participants will be implemented?

- Yes, sufficiently (1)
- Yes, **IN**sufficiently (2)
- No, and randomization is, or is likely to be, relevant (3)
- No, and randomization is, or is likely to be, **NOT** relevant (NA) (4)

Q4.12 Copy-paste the text describing **randomization** in the text box below.
 Enter "relevant" or "NA" if you chose either of the "No" options to the previous question.

________________________________________________________________

________________________________________________________________

________________________________________________________________

________________________________________________________________

________________________________________________________________

Q4.13 Does the document describe who will be **blinded** to group assignment (e.g., participants, experimenters, outcome assesors, data analysts, investigators, or any other entities)? 
Note, clinicaltrials.gov uses the term "masking" instead of blinding.

- Yes, sufficiently (1)
- Yes, **IN**sufficiently (2)
- No, and blinding is, or is likely to be, relevant (3)
- No, and blinding is, or is likely to be, **NOT** relevant (NA) (4)

Q4.14 Copy-paste the text describing **blinding** in the text box below.
 Enter "relevant" or "NA" if you chose either of the "No" options to the previous question.

________________________________________________________________

________________________________________________________________

________________________________________________________________

________________________________________________________________

________________________________________________________________

Q4.15 Does the document describe data collection methods requiring substantial **preprocessing of the data** (e.g., EEG, MRI, MEG, (molecular) genetic measures, physiological measures, hormonal measures, blood readings, or other data-intensive measures)?

- Yes, sufficiently (1)
- Yes, **IN**sufficiently (2)
- No, and preprocessing is, or is likely to be, relevant (3)
- No, and preprocessing is, or is likely to be, **NOT** relevant (NA) (4)

Q4.16 Copy-paste the text describing **preprocessing** in the text box below.
 Enter "relevant" or "NA" if you chose either of the "No" options to the previous question.

________________________________________________________________

________________________________________________________________

________________________________________________________________

________________________________________________________________

________________________________________________________________

**End of Block: Analyses**

**Start of Block: Model**

Q5.1 Does the document describe a **statistical model(s) or analysis** to assess the outcome measure(s) you identified as "**primary**" earlier in this Qualtrics form?
 For example, this could be a t-test, ANOVA, logistic regression, linear regression, multilevel regression, loglinear analysis, or any other quantitative analysis. This may also be a qualitative analyses or descriptive statistics.

- Yes, sufficiently (1)
- Yes, **IN**sufficiently (2)
- No, and a statistical model or analysis is, or is likely to be, relevant (3)
- No, and a statistical model or analysis is, or is likely to be, **NOT** relevant (NA) (4)

*Skip To: End of Block If Does the document describe a statistical model(s) or analysis to assess the outcome measure(s) yo... = No, and a statistical model or analysis is, or is likely to be, <u><strong>NOT</strong></u> relevant (NA)*

Q5.2 Copy-paste the text describing the **statistical model(s) / analysis** to test the **primary outcome measure(s)**. 
 Enter "relevant" or "NA" if you chose either of the "No" options to the previous question.

________________________________________________________________

________________________________________________________________

________________________________________________________________

________________________________________________________________

________________________________________________________________

Q5.3 Does the document describe how the authors will check for **violations of statistical assumptions** and what they are going to do with violations.
 (i.e., does the text clarify which assumptions are checked (e.g., normality, homoscedascity, linearity, homogeneity of variances, sphericity), how these assumptions are checked (e.g., type of test like Levene’s test, alpha level etc.), and what is done in cases of violations (e.g., transformations, non-parametric tests, etc.)?

- Yes, sufficiently (1)
- Yes, **IN**sufficiently (2)
- No, and a violations of statistical assumptions are, or are likely to be, relevant (3)
- No, and a violations of statistical assumptions are, or are likely to be, **NOT** relevant (NA) (4)

Q5.4 Copy-paste the text describing **violations of statistical assumptions** related to the main statistical model(s) in the text box below.
 Enter "relevant" or "NA" if you chose either of the "No" options to the previous question.

________________________________________________________________

________________________________________________________________

________________________________________________________________

________________________________________________________________

________________________________________________________________

Q5.5 Does the document describe the **inferential criteria** that will be used to make claims of an interesting or "significant" effect for the statistical model(s) / analysis for the primary outcome measures (e.g., alpha level)?

- Yes, sufficiently (1)
- Yes, **IN**sufficiently (e.g., the text fails to report the sidedness of the test) (2)
- No, and inference criteria are, or are likely to be, relevant (3)
- No, and inference criteria are, or are likely to be, **NOT** relevant (NA) (4)
- No, the study is concerned with estimation, not inference (NA) (5)

Q5.6 Copy-paste the text describing the **inferential criteria** in the text box below.
 Enter "relevant" or "NA" if you chose either of the "No" options to the previous question.

________________________________________________________________

________________________________________________________________

________________________________________________________________

________________________________________________________________

________________________________________________________________

Q5.7 Does the text specify one or more additional statistical model or analysis?

- yes (1)
- no (2)

*Skip To: End of Block If Does the text specify one or more additional statistical model or analysis? = no*

Q5.8
Does the document describe the **statistical model(s) or analysis** that will be used to test one or more of the outcome measures you identified as **"secondary"** earlier in this Qualtrics form? 
For example, this could be a t-test, ANOVA, logistic regression, linear regression, multilevel regression, loglinear analysis, or any other quantitative analysis. This may also be a qualitative analyses or descriptive statistics.

- Yes, sufficiently (1)
- Yes, **IN**sufficiently (2)
- No, and a statistical model or analysis is, or is likely to be, relevant (3)
- No, and a statistical model or analysis is, or is likely to be, **NOT** relevant (NA) (5)

Q5.9 Copy-paste the text describing the **statistical model(s) or analyses** to test one or more secondary outcome in the text box below. Enter "relevant" or "NA" if you chose either of the "No" options to the previous question.

________________________________________________________________

________________________________________________________________

________________________________________________________________

________________________________________________________________

________________________________________________________________

**End of Block: Model**

**Start of Block: Prereg badge**

*Display This Question:*

*If Which document are you reviewing? = submitted manuscript*

Q6.1 To the best of your knowledge, does the manuscript meet the four criteria for awarding a **preregistration badge** 
 (extracted from https://osf.io/tvyxz/wiki/1.%20View%20the%20Badges/ on 24 Mar 2021). This should be inferable based on the answers you provided for this form in regards to the preregistration and the manuscript.

 
1. A public date-time stamped registration is in an institutional registration system (e.g., ClinicalTrials.gov, Open Science Framework, AEA Registry, EGAP).
2. Registration pre-dates the intervention.
3. Registered design corresponds directly to reported design.
4. Full disclosure of results in accordance with registered plan.

The preregistration badge can include none, one, or both of the modifiers below.
 
For the "**Preregistered + Analysis Plan**", item 3 is worded:
3. Registered design and analysis plan corresponds directly to reported design and analysis.
 
For the "**Data Exists**" modifier, item 2 is worded:
2. Registration postdates realization of the outcomes, but the authors have yet to inspect or analyze the outcomes.
 
The manuscript could receive the following badge:

- Preregistered (2)
- Preregistetered + Analysis Plan (5)
- Preregistered (Data Exists) (6)
- None (7)

*Display This Question:*

*If Which document are you reviewing? = submitted manuscript*

Q6.2 If you answered "None" to the previous question, explain why here. Otherwise, enter "NA" in the text box below.

________________________________________________________________

**End of Block: Prereg badge**

**Start of Block: Additional comments**

Q7.1 Please add any additional comments you might have related to any part of the document you are reviewing or in relation to this form.

________________________________________________________________

________________________________________________________________

________________________________________________________________

________________________________________________________________

________________________________________________________________

**End of Block: Additional comments**

**Supplementary Material D.** Instructions for the original discrepancy review process, including our operational definitions of major, minor, and negligible issues.

**Instruction for performing discrepancy review**

Updated: 19 Apr 2021; written by Robert Thibault

**Before performing discrepancy review:**

Familiarize yourself with the project by reading [the preregistration.](https://osf.io/2bgk7/) You may also choose to read the other materials on [this project’s osf page.](https://osf.io/v4tr6/) Note, file #6 is just a word version of the Qualtrics form; file #7 is a mess and will be easier to understand when you do your own discrepancy review, so don’t waste time trying to understand it yet. File #8 and #9 are a template and example of a structured discrepancy report. After performing several discrepancy reviews, I now prefer to prepare more concise reports.

**Are you piloting discrepancy review?**

If yes, you could use the same [manuscript](https://journals.sagepub.com/doi/abs/10.1177/0956797619864598) and [preregistration](https://aspredicted.org/8vf8s.pdf) we use for our example on our project’s osf page. Alternatively, you could select any other published paper with a preregistration that you would be interested in piloting this procedure on. (Note, in our study, we will be doing this process on submitted manuscripts, not published manuscripts. We are just using a published manuscript to get familiar with the process).

-------------------------------------------------------------------------------------------

**Step-by-step instruction on how to perform discrepancy review**

1. Complete [this Qualtrics form](https://bristolexppsych.eu.qualtrics.com/jfe/form/SV_8ANnerXQEzXcJme) based on the **preregistration**. I hope the form is largely self-explanatory. However, there is surely room to improve it and to help make it applicable to a broader range of preregistrations and manuscripts. So please share constructive comments with me if you have any. If the manuscript and preregistration are substantially different, such that you wouldn’t consider them the same study, the Qualtrics form will end the survey when you enter this information.
2. Open a new instance of [the Qualtrics form](https://bristolexppsych.eu.qualtrics.com/jfe/form/SV_8ANnerXQEzXcJme) and complete it based on the [**submitted manuscript**.](https://journals.sagepub.com/doi/abs/10.1177/0956797619864598)
3. The responses from the Qualtrics forms will be sent to my Qualtrics account. Email me and I will forward you the Qualtrics output.
   i. Click on the Project ‘Standardized discrepancy form – **210324**’ (which is in the folder ‘discrepancy review’)
   ii. Click Data & Analysis --> Export & Import --> Download as TSV
   iii. Open the spreadsheet. The two bottom lines are likely the responses that you just completed.
   iv. Cut out these two rows --> open a new sheet --> paste them into columns (i.e., using the *paste transpose* function in excel).

4. Once you have the Qualtrics data, open [this google sheet](https://docs.google.com/spreadsheets/d/1wleteWGga5Dn2M_mthlydIjNyFFklTo9Oi0povZNNUY/edit?usp=sharing). Make a copy of it and rename it “discrepancyReview_[MANUSCRIPT-ID]_[YOUR-INITIALS].
   i. Paste the two columns you copied from the Qualtrics form into columns C-D in the google sheet. This action will auto-populate Columns E-G, and some NA cells in columns H-I.
   ii. Column H will auto-populate with NA if either C *or* D == 0. If column H is not auto-populated, enter ‘n’ or ‘y’. Column I will auto-populate with NA if C *and* D == 0 *or* H == ‘n’. If column I is not auto-populated, enter ‘n’ or ‘y’. In other words, if a cell is auto-populated, you don’t need to modify it.
   iii. You may manually override any cell in this google sheet if you think it is wrong (for example, you made a mistake in your coding of the Qualtrics form).
   iv. You likely want to “wrap” columns C and D to make this process visually easier.
   v. Work your way down the google sheet and fill in the red boxes (Columns H-K). Appendix A, included at the end of this document provides the definitions required to complete column J. All red boxes (except the additional comments boxes) must be filled in. These boxes will provide the data for our quantitative analyses.
   vi. The questions on preprocessing and statistical models may require expertise that you may not have. Check for overarching and obvious discrepancies (e.g., a discrepancy in the statistical test used). If you feel these items are too far beyond your expertise, type “unable to assess” in columns H-J for that item. This option can be used for other items, but try your best to only use it when necessary.
   vii. Note, we only consider content discrepant (“y” to column H) if the content is different. If the manuscript includes a reproducible definition and the prereg has a vague definition, but they are compatible, then enter “n” in this column.

5. Open a template [*discrepancy report*](https://drive.google.com/file/d/15zXMEQfTjnvlp4XVqGm-jguW15Yo71GT/view?usp=sharing). Make a copy of it and rename it “discrepancyReport_[MANUSCRIPT-ID]_[YOUR-INITIALS]. ~~If you prefer, you can choose to use the~~ [~~more systematic template~~](https://drive.google.com/file/d/1D1bougTVFRLBHleGPF6mJv9ejLaFJnSA/view?usp=sharing) ~~that results in a much longer discrepancy review.~~
   i. Complete the report. Most of the information to populate this report will come from the red boxes you just completed in the google sheet.
   ii. Only include items for which you have recommendations.
   iii. Try to make this report palatable to the authors. Keep in mind, we are trying to help them improve their manuscript, we are not trying to accuse them of wrong-doing.
   iv. You may deviate from the example discrepancy reports (e.g., collapse multiple recommendations into one) if you think this will make the report more clear or concise.
   v. Writing this report is subjective. But this is not an issue for our study, because (a) if discrepancy review is used as a regular practice, it would also be subjective, and (b) as far as our study goes, the report is *procedural*. The data we will analyze comes from the google sheet (which hopefully is less subjective).
   vi. [new Apr 15] I added some boilerplate text in Appendix B of this document that you may choose to adapt to respond to some common issues that come up during discrepancy review.

WHEN YOU ARE DONE WRITING THE DISCREPANCY REPORT, PLEASE COMPLETE [THIS SHORT SURVEY](https://bristolexppsych.eu.qualtrics.com/jfe/form/SV_bNoBhTwrHKCOAxE) (it is a bit messy at the moment, but should serve its purpose nonetheless).

**After submitting the discrepancy review** (note, the rest of the items are not relevant if you are piloting discrepancy review). [At the moment, Robert is the only point of contact for the journals and their editors. Thus, you do not have to worry about the items 6-13 below. Robert will inform you if he needs your input on any of these]

6. Log the discrepancy review you performed in the discrepancy review log.
7. ~~Complete~~ [~~this short survey~~](https://bristolexppsych.eu.qualtrics.com/jfe/form/SV_2tn2nizxi84Lagl) ~~about your experience with discrepancy review. Do this after every review. If you don’t have much to say, you can leave most of the boxes empty.~~ This survey only had two questions. They have now been integrated into the discrepancy report in the section “*information for internal use*”. So you do NOT need to go to that survey link.
8. The editor may invite you to perform additional rounds of review.

**After an accept or reject decision has been made by the editor**

9. Complete [this survey](https://bristolexppsych.eu.qualtrics.com/jfe/form/SV_2fNteM2n40m0NL0) about your experience with discrepancy review.
10. Contact the action editor and ask them if they are willing to [complete this questionnaire](https://bristolexppsych.eu.qualtrics.com/jfe/form/SV_elnWtDdRqWzYizQ) about their experience.
11. If the manuscript authors have not already been contacted by the journal, send an email asking if they are willing to [complete this questionnaire](https://bristolexppsych.eu.qualtrics.com/jfe/form/SV_4GCDqPpzfsEE4No). If the authors have not yet been informed about the study, then do so in this email.
12. Finish the discrepancy review log for this entry.
13. Perform discrepancy review on the final accepted or published manuscript. Assess whether discrepancies are now disclosed or no longer present [this step may also be performed by another reviewer instead of the reviewer who wrote the discrepancy review].

**Appendix A.** Definitions for Column J: *Discrepancies and issues of transparency*. This process is somewhat subjective. We will be checking inter-rater reliability as we go.

**NA**: Not applicable. (E.g., neither the preregistration nor the manuscript mention an item and that item is irrelevant; for example, blinding and randomization in an observational study).

**None**: The item has been reported in a reproducible manner and there are no discrepancies between the preregistration and the manuscript.

**Negligible (0)**: There are slight discrepancies, additions, or omissions between the preregistration and the manuscript; AND/OR the item is not quite described in a reproducible manner. HOWEVER, this issue is unlikely to substantially affect the quality of the research and could be left unaddressed.

**Minor (1)**: There are minor discrepancies, additions, or omissions between the preregistration and the manuscript; AND/OR the item is not described in a reproducible manner or is not described at all. This issue should be addressed.

**Major (2)**: There are considerable discrepancies, additions, or omissions between the preregistration and the manuscript; AND/OR the item is not described; AND/OR the item is not described in a reproducible manner. We highly recommend this issue be addressed.

**Appendix B.** Boilerplate text you may choose to adapt for your discrepancy review report.

**1.**	**If the study presented in the registration and manuscript are not the same.**

The manuscript and the registration it points to [registration ID or URL] align only in the sample they use. Thus, I recommend the manuscript includes a statement along the lines of: “The participant sample is registered at [registration ID or URL]. However, the [outcome measures and analyses] reported in the manuscript are not included in the registration and the [intervention conditions and outcome measures] in the registration are not reported in this manuscript.”

**2.**	**If the preregistration is uploaded to the OSF HOME page, but not officially registered.**

Preregistrations should be public and permanent. The current preregistration is uploaded to an Open Science Framework (OSF) project page. This document can be removed or modified. I recommend registering this project page by following the [instructions available here](https://help.osf.io/hc/en-us/articles/360019738834-Create-a-Preregistration) and selecting “Preregister a project you already have on OSF”. This will make the preregistration permanent and public.

**3.**	**If the timing of the registration is unclear.**

I recommend that authors explicitly state at what stage of the project the registration was uploaded. For example, was it before recruiting the first participant, during participant enrolment, after enrolment but before data analysis, or after data analysis?

**4.**	**If the inclusion and exclusion criteria are extensive or hard to align or there appears to be many discrepancies.**

The inclusion/exclusion criteria do not match between the manuscript and the registration. If the full inclusion/exclusion list in the registration is correct, the manuscript could simply point readers to the registration for the full list of inclusion/exclusion criteria.

**5.**	**If many numbered hypotheses are made in the registration, but they are presented in a confusing manner in the manuscript.**

The hypotheses are difficult to compare between the preregistration and the manuscript because they are presented in different formats (i.e., itemized hypotheses versus predictions embedded in multiple pages of text). A few differences seem apparent. I recommend the authors match the hypotheses in the manuscript to those in the preregistration (e.g., by putting the preregistered hypothesis number next to the hypothesis in the manuscript). I also recommend they mention any deviations they made.

**6.**	**If items are neither reported in the manuscript nor the registration.**

To maximize transparency, several items of information that are not included in the registration or the submitted manuscript could be added to the manuscript. I recommend these appear alongside a note that this information was not preregistered. These could include

[Delete items as necessary]

- Inclusion/exclusion criteria for participants.
- Inclusion/exclusion criteria for data (e.g., outliers, failed attention check)
- What has been done with missing data
- Whether violations of statistical assumptions were tested for
- What the inferential criteria being used are (e.g., are you using p<0.05 and accounting for multiple comparisons to claim a significant effect).
- How was randomization implemented (e.g., allocation sequence)
- Was anyone blinded (e.g., participants, outcome assessors, data analysts)
- Details on data preprocessing.

**Supplementary Material E.** Instructions guiding the updated discrepancy review process.

**Instruction for performing STREAMLINED [updated] discrepancy review** [no Qualtrics forms and no spreadsheets]

Updated on 12 May 2021.

We will be testing the feasibility of two different types of discrepancy review (i) [the one we’ve been doing so far](https://docs.google.com/document/d/1_619KHFZ3dfLalibQ2759MXMmErA5hYFMEypaJMiz7M/edit?usp=sharing) (i.e. the *original* version), and (ii) this “*streamlined*” version.

They are different in a few ways:

- In the original discrepancy review method, we were collecting data to analyze for our feasibility study outcomes AND for writing the discrepancy report. In the streamlined version, we are ONLY collecting information to help write the discrepancy report. I am aiming to remove as much chaff as possible to make discrepancy review a process that journals could feasibly contract someone to perform (i.e., to make the effort worth the benefit).
- The streamlined version looks at many fewer items. Hopefully this will substantially reduce the time it takes to perform discrepancy review, but still maintain the main benefits.
- Because the streamlined version is less thorough, it can only be used as evidence that a preregistration badge should NOT be earned. It is not detailed enough to establish that a preregistration badge should be earned. Thus, journals that award preregistration badges may prefer the original version whereas journals that do not award badges may be happier with the streamlined version.

------------------------------------------------------------------------------------------------

**Step-by-step instruction on how to perform STREAMLINED discrepancy review**

(These instructions were adapted from [this Qualtrics form](https://bristolexppsych.eu.qualtrics.com/jfe/form/SV_8ANnerXQEzXcJme), which was in turn adapted from the work of Olmo van den Akker (https://osf.io/ta3yd/) and Bakker et al. (<https://psyarxiv.com/cdgyh/>).

**Ensure you have a copy of both the preregistration and the submitted manuscript.** For the purpose of this form, we define a preregistration as a publicly available document that includes elements of a study protocol and that was archived before the beginning of a study. This definition includes pre-analysis plans and prospective registrations. **Open a blank discrepancy** [**report.**](https://drive.google.com/file/d/15zXMEQfTjnvlp4XVqGm-jguW15Yo71GT/view?usp=sharing)

**Go through the numbered items below one at a time.** If you find issues, draft your comments to the authors in the discrepancy report. If there are no issues, you do not need to make any notes or write anything in the discrepancy report.

**Record the time it takes to prepare the report precisely** (i.e., set a stopwatch and pause it when you leave your desk or move to another task). Try not to feel rushed because there is a timer. At this point, we just want to know how long it takes.

**I am unsure whether it would be easier/quicker to do this streamlined version of discrepancy review with a Qualtrics form or spreadsheet. At the moment, let’s try it out by just following the instructions in this document and writing notes in a text document. Let me know if you have any thoughts on this.**

**You don’t need to write or record anything more than what will appear in the discrepancy report to authors.**

---------------------------------------------------------------------------------------------------

1. **Read the abstract of the manuscript and quickly scan the preregistration. Are the manuscript and preregistration sufficiently similar that you would consider that they reflect the same study?**

We do not yet have a strict operationalization of what qualifies as "different studies". Here is some information to help you decide:

(1) If the manuscript points to a registration, but the only thing that overlaps between the manuscript and the registration is the participant sample, then we consider these to be different studies. For example, a manuscript may report only on qualitative or demographic data that was collected during a clinical trial, but no info about the qualitative or demographic data appear in the clinical trial registry (i.e., not in the aims or the outcomes of the trial registration).

(2) Do you think it would be more appropriate for the manuscript to include:

(i) an overarching statement along the lines of "This manuscript reports a secondary/exploratory analysis of a study registered at [registration ID], but none of the outcome measures or analysis in this manuscript are included in the registration and none of the registered outcome measures or analyses are included in the manuscript"; OR

(ii) a disclosure for every discrepancy between the registration and manuscript (this could be in the manuscript text, or in an appendix that outlines all the discrepancies).

If you think (i) is more appropriate, then we consider these to be "different studies".

If you consider the preregistration and manuscript to be different studies, you do not need to perform discrepancy review. Instead, write a few sentences that you would suggest the authors include in their manuscript. Here is an example comment:

*The manuscript and the registration it points to [registration ID or URL] align only in the sample they use. Thus, I recommend the manuscript includes a statement along the lines of: “The participant sample is registered at [registration ID or URL]. However, the [outcome measures and analyses] reported in the manuscript are not included in the registration and the [intervention conditions and outcome measures] in the registration are not reported in this manuscript.”*

2. **Does the preregistration meet the criteria for a preregistration?**

In other words, is it permanent, read only, time-stamped, public, non-ambiguously accessible, and in a third party repository? A registration meets these criteria if it is stored at one of the following website, (e.g., clinicaltrials.gov, OSF Registries, AsPredicted, PROSPERO, EUCTR, ANZCTR, DRKS, ISRCTN, ICRTP). If the registration is stored at the OSF HOME page, use or adapt the following text:

*Preregistrations should be public and permanent. The current preregistration is uploaded to an Open Science Framework (OSF) project page. This document can be removed or modified. I recommend registering this project page by following the* [*instructions available here*](https://help.osf.io/hc/en-us/articles/360019738834-Create-a-Preregistration) *and selecting “Preregister a project you already have on OSF”. This will make the preregistration permanent and public.*

In some cases, authors may upload a preregistration without any author names for a blinded review process. In this case, the document may not be permanent or public because it is only being used for peer review. If you believe this to be the case, you could ask the authors to confirm that their preregistration is uploaded to a permanent and public page (e.g. OSF Registries).

3. **Is the registration date questionably close to the submission date?**

Such that information in the preregistration may have been documented after the study began? The date range that could be considered questionable will vary depending on the type of study (e.g., a data collection period of two years versus an online survey of 100 participants). If yes, include an adaptation of the following text:

*I recommend the authors explicitly state at what stage of the project the registration was made. For example, was it before recruiting the first participant, during enrolment, after enrolment but before data analysis, or after data analysis?*

**NOTE, for questions #4-8**, we are only interested in **undisclosed** discrepancies. If a discrepancy exists, but disclosed, you don’t need to write anything about it in your discrepancy report.

4. Identify all the **hypotheses / aims / goals / research questions** from the preregistration and the manuscript. Do they differ (e.g., in number, content, or prioritization of “primary” and “secondary”).

5. Identify the **independent variable(s), exposure variable(s), or study/experimental grouping** (these can be manipulated or non-manipulated).
   a. Do the number of groups/arms match?
   b. Does the description of the groups match between the documents?

For this question, do**n’t** spend time looking for minor discrepancies between the registration and the manuscript.

6. Identify the **outcome measures, dependent variables, or endpoint measures**?
   a. Do they differ between the preregistration and the manuscript (e.g., in number, content, or prioritization).

For the manuscript, extract the outcomes from the Results section rather than the Methods section. We advise this because sometimes a manuscript will mention outcomes in the methods section, but never report them in the results section, and vice versa.

7. Identify the **analyses** in the registration and the manuscript. These could be inferential, descriptive, or qualitative.
   a. Are there the same number of analyses in the registration and the manuscript?
   b. Are there any major discrepancies in analyses that appear in both the registration and manuscript (e.g., a t-test turned into an ANOVA)?

For this question, do**n’t** spend time looking for minor discrepancies between analyses in the manuscript and registration. If the registration doesn’t include any analysis plans (as is the case for most clinicaltrials.gov registrations), you could use the following text:

*I recommend the authors explicitly state that analysis plans were not registered.*

8. If you have concerns about discrepancies or questionable research practices for any of the items below, you may raise them. Don’t spend too much time looking for these, just note them if you notice a reason for concern when going through the questions 4-7 above.

*·	sample size*

*·	control variables / covariates / moderators*

*·	Inclusion/exclusion criteria for participants*

*·	Inclusion/exclusion criteria for data (e.g., outliers, failed attention check)*

*·	What has been done with missing data*

*·	Whether violations of statistical assumptions were tested for*

*·	What the inferential criteria being used are (e.g., are you using p<0.05 and accounting for multiple comparisons to claim a significant effect).*

*·	How was randomization implemented (e.g., allocation sequence)*

*·	Was anyone blinded (e.g., participants, outcome assessors, data analysts)*

*·	Details on data preprocessing.*

WHEN YOU ARE DONE WRITING THE DISCREPANCY REPORT, PLEASE COMPLETE [THIS SHORT SURVEY](https://bristolexppsych.eu.qualtrics.com/jfe/form/SV_bNoBhTwrHKCOAxE) (it is a bit messy at the moment, but should serve its purpose nonetheless).

**Supplementary Material F.** Qualtrics form of follow-up questions after discrepancy review, including operational definitions of the importance of addressing discrepancies.

**Discrepancy review follow-up Qs**

**Survey Flow**

**Block: Default Question Block (8 Questions)**

| Page Break |
| --- |

**Start of Block: Default Question Block**

Q7 Enter the manuscript ID (e.g., NTR-2021-000)

________________________________________________________________

Q6 Unless you just completed a discrepancy report, skip this question. Otherwise, how many **minutes** did it take you to prepare the discrepancy report (not including this survey)?

________________________________________________________________

Q1 Select the boxes if the registration includes that item (i.e., does it meet the ICMJE registration criteria)?

- It names the intervention, independent variables, exposure variables, or other grouping (1)
- Participant inclusion and/or exclusion criteria (2)
- Target sample size (For meta-analyses, leave this box blank, it doesn't apply) (3)
- Outcome. (If the registration is on clinicaltrials.gov or another clinical trial registry, there MUST be at least one outcome labelled as primary and that outcome must include the metric used and the timeframe—e.g., Hamilton Anxiety Rating Scale at 3 months post treatment). (4)

Q2 Select the option below that is most appropriate.

- The manuscript and registration have no relation. The manuscript should not mention the registration. (1)
- The manuscript reports a non-registered secondary or exploratory analysis of the registered study sample. In other words, the participant sample and the intervention, independent variables, or exposure variables may match between the registry and the manuscript, but the outcome measures and analysis have no relation to the registered outcome measures and analysis. The manuscript should include the registration ID or DOI and also state that it reports outcome measures and analyses that were not prospectively registered. (2)
- The manuscript and registration reflect the same study and there is at least one undisclosed discrepancy. (3)
- The manuscript and registration reflect the same study and there are no discrepancies, or all discrepancies are disclosed. (4)

*Display This Question:*

*If Q2 = The manuscript and registration reflect the same study and there is at least one undisclosed discrepancy.*

Q3 **How important do you think it is that these discrepancies are addressed?** You may interpret this question in several ways, including (but not limited) to the following:

1. **Risk** **of bias.** Do the discrepancies suggest there is a high risk of bias in the design, conduct, or reporting of the study that could lead to over- or under-estimates of the effect under investigation?

2. **Questionable research practices.** Do the discrepancies suggest that questionable research practices such as p-hacking, optional stopping, or post-hoc data exclusion may have occurred?

3. **Transparency and** **Reproducibility.** Do the discrepancies impact the transparency of the manuscript? Would it be difficult for someone to reproduce the study and/or analyses because of discrepancies?

4. **Truth of the findings.** Do the discrepancies make you feel the results are less likely to be true (i.e., correctly report on the phenomenon of interest that exists in the world)?

- Quite important (1)
- Somewhat important (2)
- Not important (3)

Q4 How confident are you regarding the previous question.

- very confident (1)
- confident (2)
- somewhat confident (3)
- hardly more confident than chance (i.e., randomly selecting one of the options) (4)
- no more confident than chance (i.e., randomly selecting one of the options) (5)

Q8 Briefly, can you provide the reasons for your answers to the previous two questions.

________________________________________________________________

________________________________________________________________

________________________________________________________________

________________________________________________________________

________________________________________________________________

Q5 Please provide any thoughts you might have about this project, preregistration, or discrepancy review in general. Don't be shy!

________________________________________________________________

________________________________________________________________

________________________________________________________________

________________________________________________________________

________________________________________________________________

**End of Block: Default Question Block**

**Supplementary Material G.** Power analysis and precision analysis code.

library(statpsych) # for the precision analysis

\#### documentation available at : https://csass.ucsc.edu/powerprecision.pdf

a = .05 # alpha value

p1= .1 # proportion with at least one primary discrepancy in the control group

p2= .33 # proportion with at least one primary discrepancy in the experimental group

w = .20 # precision

\# n per group for inputs above for POWER analysis

nPower <- power.prop.test(n = NULL, p1, p2, sig.level = .05, power = .80, alternative = "two.sided")

nPower <- nPower$n %>% round(0)

\# n per group for inputs above for PRECISION analysis

nPrecision <- size.ci.prop2(a, p1, p2, w)
